# Deep Learning–Based Multiclass Classification of Mitral Valve Etiologies Using Limited B-Mode and Color Doppler Echocardiography: Internal and External Validation

**DOI:** 10.64898/2026.01.09.26343662

**Authors:** Dawun Jeong, Moon-Seung Soh, Jaeik Jeon, Hyunseok Jeong, JunHeum Cho, Jiyeon Kim, Jina Lee, Seung-Ah Lee, Joon-Han Shin, Yeonggul Jang, Yeonyee E. Yoon, Hyuk-Jae Chang

**Author notes:** The first two authors contributed equally to this work. **Corresponding authors:** Yeonyee E. Yoon, MD, PhD Professor, Division of Cardiology, Cardiovascular Center Seoul National University Bundang Hospital 82, 173 Beon-gil, Gumi-ro, Bundang-gu, Seongnam-si, Gyeonggi-do, Republic of Korea And Yeonggul Jang, PhD Ontact Health Co., Ltd., Seoul, Republic of Korea 50-5, Ewhayeodae-gil, Seodaemun-gu, Seoul, Republic of Korea.

## Abstract

**Background:** Accurate etiologic classification of the mitral valve (MV) is essential for guiding clinical management but remains dependent on expert visual interpretation. Despite advances in artificial intelligence (AI)-based quantitative analysis, automated morphologic interpretation under routine imaging conditions remains limited.

**Objectives:** To develop and validate a deep learning (DL) framework for multiclass classification for major MV etiologies using limited routine transthoracic echocardiography (TTE) views.

**Methods:** A multi-view DL model was developed to classify five MV etiologies (normal, rheumatic, degenerative, prolapse, and functional). The developmental dataset comprised 4,344 TTE examinations from a nationwide multicenter registry. Validation was performed using an internal test dataset and an independent external test dataset (2,262 TTE examinations). Prespecified subgroup analyses were conducted according to mitral regurgitation (MR) severity and automated image quality (IQ).

**Results:** The model demonstrated robust performance across all MV etiologies in both internal and external datasets. In the internal test dataset, area under the receiver operating characteristic curve (AUROC) values ranged from 0.968 to 0.997 across etiologies, with higher performance observed for normal valves and rheumatic disease. In the external test dataset, discriminative performance remained preserved (AUROC, 0.931-0.992), despite differences in disease distribution and MR severity. Sensitivity for MV prolapse increased markedly with moderate or greater MR compared with mild MR, whereas degenerative disease showed persistently lower sensitivity across MR severity. Diagnostic performance remained stable across IQ strata, with comparable accuracy and macro-F1 scores in all-adequate and partially suboptimal examinations. In post-hoc analyses of cases with multiple MV etiologies, the model correctly identified at least one expert-assigned etiology in 85.7% of cases.

**Conclusions:** DL-based analysis of limited, routinely acquired TTE views enables reliable multiclass classification of MV etiologies. This approach may complement quantitative automation and expert visual assessment, supporting more consistent and scalable MV evaluation in routine echocardiographic practice.

## INTRODUCTION

Echocardiography is the primary imaging modality for evaluating valvular heart disease (VHD). Comprehensive echocardiographic assessment of VHD requires both morphological and functional evaluation of the affected valve.^1,2^ Quantitative hemodynamic measurements, such as Doppler-derived transvalvular gradients and flow parameters, play a central role in determining functional severity and have already been standardized to a large extent.^1,2^ In contrast, the assessment of morphological abnormalities that underlie valvular dysfunction still relies heavily on expert visual interpretation. This process is inherently operator-dependent, experience-driven, and therefore less reproducible across readers and institutions.

Artificial intelligence (AI)-based interpretation of echocardiography has rapidly expanded in recent years.^3,4^ Initial efforts mainly focused on automating quantitative measurements, and many manual parameters can now be reliably extracted by AI.^4–8^ These advances have also been applied to VHD, where AI can automate key components of diagnosis and grading.^6,9–11^ However, the evaluation of valvular morphology remains largely dependent on human expertise. This challenge is particularly pronounced for the mitral valve (MV), where comprehensive assessment requires detailed evaluation of not only the leaflets but also the subvalvular apparatus. Moreover, determining the underlying etiology of MV disease is essential for guiding management decisions,^1,2^ yet accurate classification is difficult outside specialized centers. These limitations underscore the need for AI systems capable of consistently providing expert-level morphological interpretation.

Therefore, we sought to develop a deep learning (DL) model capable of MV etiology classification using limited B-mode and color Doppler echocardiographic inputs. The model was designed to differentiate normal valves from four major etiologic categories of MV disease – rheumatic, degenerative, prolapse, and functional – encompassing both mitral stenosis (MS) and mitral regurgitation (MR). We further aimed to rigorously evaluate model performance using internal and external test datasets from independent cohorts to assess its generalizability and clinical applicability. An overview of the study design, multi-view DL framework, development and validation cohorts, and etiology-specific model performance is provided in **Central Illustration**.

## MATERIAL AND METHODS

### Study Population

In this study, the DL framework was developed using data from the Open AI Dataset Project (AI-Hub), a nationwide echocardiography repository funded by the Ministry of Science and ICT of the Republic of Korea.^12^ The database includes approximately 30,000 transthoracic echocardiographic (TTE) examinations, obtained retrospectively from several tertiary medical centers across South Korea between 2012 and 2021, representing a diverse range of cardiovascular pathologies.^6–8,13^ Rather than consisting of consecutive TTE examinations collected from a single institution or during a fixed period, the AI-Hub database was constructed to include normal studies and a broad spectrum of cardiovascular disease across multiple centers. For model development, a total of 4,344 TTE examinations from 4,317 patients were used, comprising both cases with MV pathology and individuals without MV disease. Normal controls were identified from the “normal” category of the database and consisted of asymptomatic individuals who underwent TTE examinations as a part of routine health evaluation. Only examinations demonstrating entirely normal MV structure and function were included in the control group. Cases with MV disease were obtained from the “MV disease” category and identified using ICD-10 diagnostic codes I05.0, I05.1, I34.0, and I34.2. TTE examinations demonstrating the presence of MS or MR of at least mild severity were included, whereas studies confirming normal MV function were excluded.^1,2,14^ Examinations where assessment of the native MV could be confounded by prior surgery-related materials or structural alterations were excluded. These included cases with any prosthetic heart valve, prior MV surgery or intervention (including percutaneous mitral valvotomy), tricuspid annuloplasty, ascending aorta replacement, or complex congenital heart disease. In contrast, procedures that do not directly affect the native MV, such as pacemaker or cardioverter-defibrillator implantation, cardiac resynchronization therapy, simple atrial or ventricular septal defect repair, coronary artery bypass grafting, or abdominal aorta aneurysm repair, were not considered exclusion criteria. All eligible examinations were subsequently categorized by underlying MV etiology. Cases with multiple MV disease etiologies were excluded from the developmental dataset. The final developmental dataset consisted of 1,391 normal studies, 469 rheumatic cases, 162 degenerative cases, 1,478 prolapse cases, and 844 functional cases. This cohort was then randomly divided into training, validation, and internal test subsets in an 8:1:1 ratio, yielding 3,458 patients in the training set, 429 in the validation set, and 430 in the internal test set, corresponding to 3,485, 429, and 430 TTE examinations, respectively. (**Table 1**) To avoid data leakage, all TTE examinations from patients who underwent multiple TTE studies were assigned exclusively to the training set. In contrast, patients included in the validation and internal test set each contributed a single TTE examination.

**Table 1.**
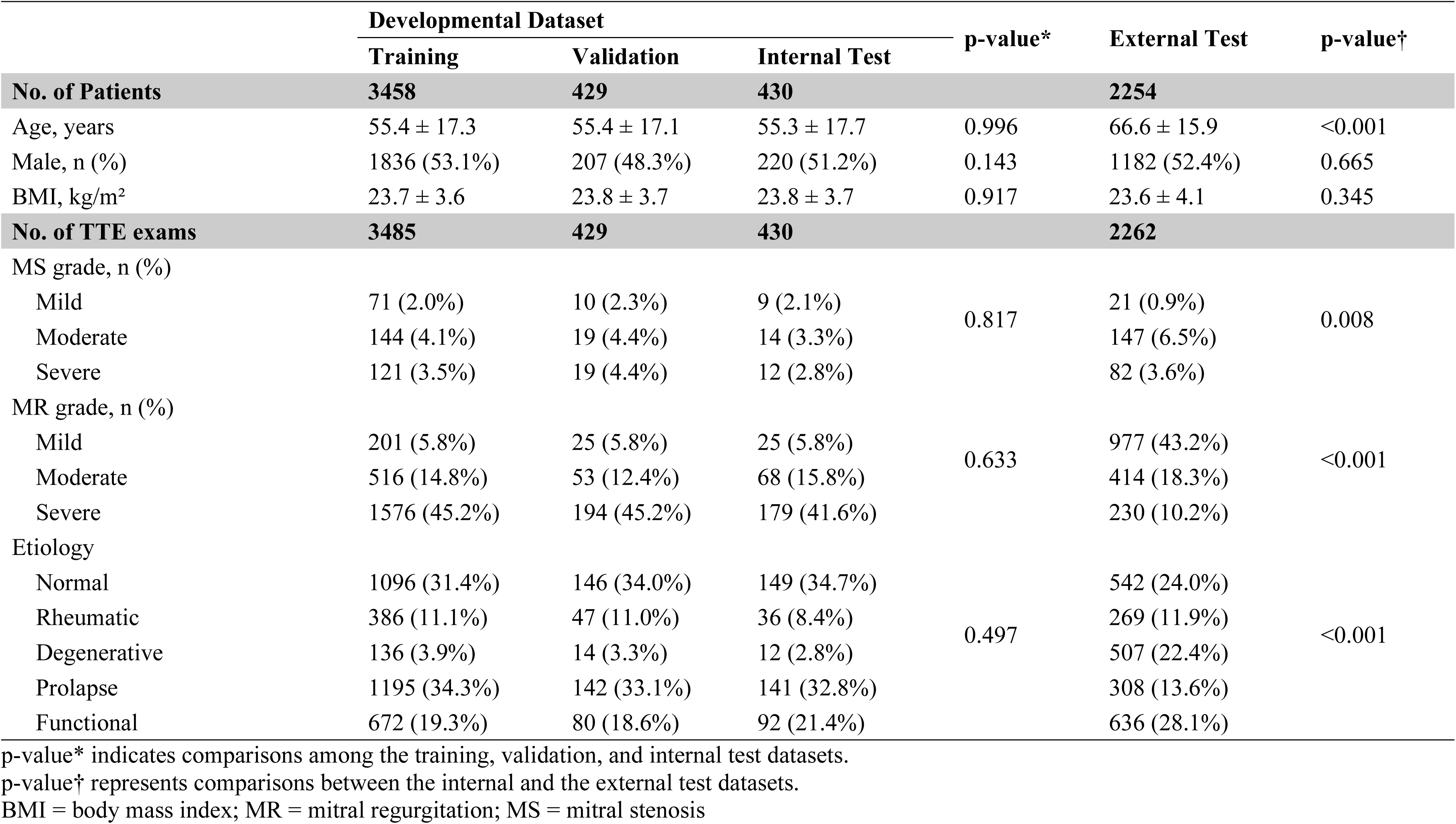
Baseline Characteristics.

|  | Developmental Dataset |  |  | p-value* | External Test | p-value† |
| --- | --- | --- | --- | --- | --- | --- |
|  | Training | Validation | Internal Test |  |  |  |
| **No. of Patients** | **3458** | **429** | **430** |  | **2254** |  |
| Age, years | 55.4 ± 17.3 | 55.4 ± 17.1 | 55.3 ± 17.7 | 0.996 | 66.6 ± 15.9 | <0.001 |
| Male, n (%) | 1836 (53.1%) | 207 (48.3%) | 220 (51.2%) | 0.143 | 1182 (52.4%) | 0.665 |
| BMI, kg/m<sup>2</sup> | 23.7 ± 3.6 | 23.8 ± 3.7 | 23.8 ± 3.7 | 0.917 | 23.6 ± 4.1 | 0.345 |
| **No. of TTE exams** | **3485** | **429** | **430** |  | **2262** |  |
| MS grade, n (%) |  |  |  |  |  |  |
| Mild | 71 (2.0%) | 10 (2.3%) | 9 (2.1%) | 0.817 | 21 (0.9%) | 0.008 |
| Moderate | 144 (4.1%) | 19 (4.4%) | 14 (3.3%) |  | 147 (6.5%) |  |
| Severe | 121 (3.5%) | 19 (4.4%) | 12 (2.8%) |  | 82 (3.6%) |  |
| MR grade, n (%) |  |  |  |  |  |  |
| Mild | 201 (5.8%) | 25 (5.8%) | 25 (5.8%) | 0.633 | 977 (43.2%) | <0.001 |
| Moderate | 516 (14.8%) | 53 (12.4%) | 68 (15.8%) |  | 414 (18.3%) |  |
| Severe | 1576 (45.2%) | 194 (45.2%) | 179 (41.6%) |  | 230 (10.2%) |  |
| Etiology |  |  |  |  |  |  |
| Normal | 1096 (31.4%) | 146 (34.0%) | 149 (34.7%) | 0.497 | 542 (24.0%) | <0.001 |
| Rheumatic | 386 (11.1%) | 47 (11.0%) | 36 (8.4%) |  | 269 (11.9%) |  |
| Degenerative | 136 (3.9%) | 14 (3.3%) | 12 (2.8%) |  | 507 (22.4%) |  |
| Prolapse | 1195 (34.3%) | 142 (33.1%) | 141 (32.8%) |  | 308 (13.6%) |  |
| Functional | 672 (19.3%) | 80 (18.6%) | 92 (21.4%) |  | 636 (28.1%) |  |

p-value\* indicates comparisons among the training, validation, and internal test datasets.
p-value† represents comparisons between the internal and the external test datasets.
BMI = body mass index; MR = mitral regurgitation; MS = mitral stenosis

An external test cohort was additionally assembled using 2,262 TTE examinations from 2,254 patients obtained at Ajou University Hospital between 2020 and 2024. Normal controls in this cohort were randomly selected from subjects whose studies were reviewed and confirmed to demonstrate normal MV morphology and function. Cases representing a broad spectrum of MV etiologies were included to ensure comprehensive evaluation of model performance. The external dataset comprised the following etiologic categories: normal (n=542), rheumatic (n=269), degenerative (n=507), prolapse (n=308), and functional (n=636). The institutional review boards of all participating hospitals approved the study protocols and waived the requirement for informed consent owing to the retrospective and observational study design (2021-0147-003 / CNUH 2021-04-032 / HYUH 2021-03-026-003 / SCHBC 2021-03-007-001 / B 2104/677-004). Additional approval for the construction and use of the external test cohort was obtained from the IRB of Ajou University Hospital (AJOUIRB-DB-2024-430). All clinical and echocardiographic data were fully anonymized prior to analysis.

### TTE Acquisition and Utilization

All TTE examinations were performed by trained sonographers or cardiologists and were initially interpreted by board-certified cardiologists specializing in echocardiography, following current clinical practice guidelines.^1,2,15^ To ensure accurate labeling of the developmental dataset, including the training, validation, and internal test subsets, two expert cardiologists (S.A.L., > 10 years of experience; Y.E.Y., > 15 years of experience) independently reviewed the complete TTE studies for each case. Any discrepancies were resolved through consensus, and final diagnostic labels were assigned accordingly.

Standardized echocardiographic definitions were applied for MV etiologies on the basis of valve morphology and pathophysiologic mechanism. MV prolapse was defined as systolic displacement of one or both leaflets ≥2 mm beyond the annular plane and included both Barlow disease and fibroelastic deficiency phenotypes.^16^ Rheumatic etiology was defined by characteristic rheumatic features, including leaflet thickening predominantly at the tips, commissural fusion, chordal involvement, and restricted leaflet motion.^14^ Functional etiology was defined as mitral regurgitation occurring in the setting of structurally normal leaflets, secondary to ventricular or atrial remodeling.^17^ Degenerative etiology was defined as non-rheumatic calcific degenerative MV disease, characterized primarily by definite mitral annular calcification with corresponding degenerative valvular morphology, in the absence of criteria for prolapse or rheumatic disease.^1^

Importantly, although expert annotations were derived from a comprehensive review of the entire TTE examination, only a limited set of standard views was used as input to the DL framework. The selected views included the parasternal long-axis (PLAX) and apical four-chamber (A4C) views, with and without color Doppler. Because the present framework was designed to classify a single dominant etiology of MV pathology, cases with overlapping morphological features (e.g., degenerative changes with superimposed functional regurgitation) were excluded from the developmental dataset. These cases were retained separately and used for additional analyses to further assess model robustness.

For the external test dataset, the same standardized definition of MV etiology was used; however, labeling procedures were designed to reflect real-world clinical practice rather than expert-adjudicated reannotation of the full cohort. The etiologies of MV pathology were primarily determined from the original TTE reports generated at the contributing institution. All cases in the external test dataset were assigned a single predominant MV etiology, which was independently reviewed and confirmed by an experienced cardiologist (M.S.S., > 5 years of experience) who was not involved in labeling the development dataset. In case of overlapping morphologic features, the predominant etiology was assigned according to the principal echocardiographic mechanism judged to best explain the overall MV pathology, consistent with guideline-based echocardiographic assessment of valvular etiology and mechanism.^2,14,16,17^

To preserve real-world clinical variability, both the developmental and external test datasets included all available TTE examinations without exclusion based on image quality (IQ) or examination completeness. Examinations were retained even when only the PLAX view was available, as well as when specific target views were missing, or image quality was suboptimal. When multiple video clips were available for a given TTE view, all clips were provided to the DL-based model as input. Conversely, the absence of any individual view did not preclude inclusion of the examination. IQ was assessed using an automated, model-based approach rather than subjective human visual grading.

### DL-Based Framework

To select the target TTE views and assess IQ, we applied our previously developed AI-based automatic TTE analysis system (Sonix Health; Ontact Health Co., Ltd., Korea).^5–8,18^ Target views included the PLAX view, a PLAX view with zoomed visualization of the MV, both acquired with and without color Doppler, and the A4C view with and without color Doppler. These target views were initially identified using the system’s view classification module, after which investigators then reviewed the selected clips to remove misclassified views and recover any missing ones, thereby ensuring consistent and accurate inputs for the DL model. Although the underlying view classification algorithm was developed across a broader range of echocardiographic imaging modes, only the B-mode and color Doppler view categories relevant to the present framework were used in this study. Detailed target view categories and classifier performance are provided in **Supplementary Methods 1**. Following view selection, the IQ of the retained video clips was evaluated using the system’s segmentation module.^5–8,18^ IQ was quantified from the entropy of the segmentation outputs and categorized into four levels (good, fair, poor, and non-diagnostic). Detailed descriptions of the segmentation-based IQ assessment are provided in **Supplementary Methods 2**.

Building upon the selected TTE views, we developed a multi-view DL architecture for MV disease etiology classification, designed to reflect the established clinical assessment workflow. The proposed architecture consists of three key components: (1) a shared MViT-Small encoder^19^ that extracts unified spatiotemporal representations across all input views; (2) view-specific projection heads that disentangle and process anatomical features while accommodating missing views via learnable embeddings; and (3) a learnable gating mechanism that dynamically refines valve-related features by integrating zoomed morphological details when available. (**Figure 1**)

**Figure 1.**
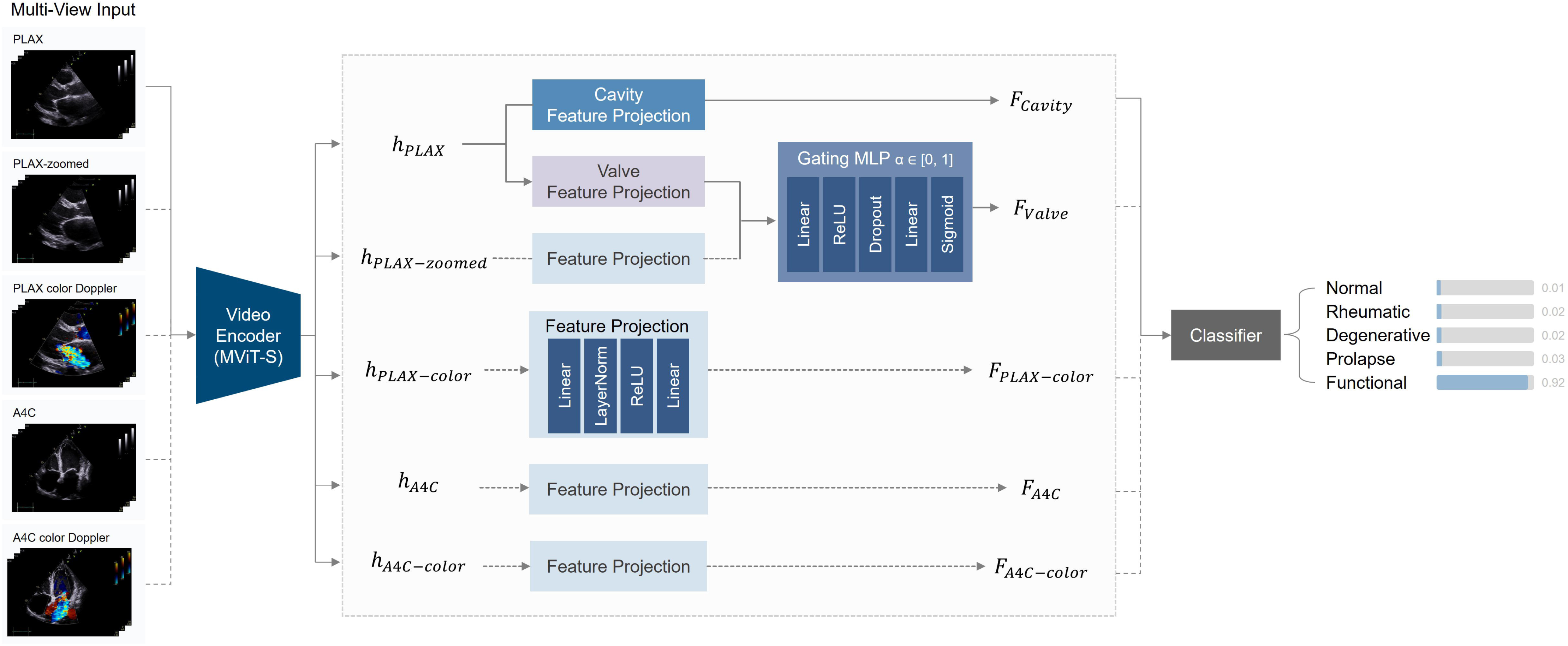
Multi-view Deep Learning Architecture for Mitral Valve Disease Etiology Classification. The model integrates limited routine echocardiographic views using a shared spatiotemporal encoder and view-specific feature projections to infer mitral valve etiology.

A shared spatiotemporal encoder based on MViT-Small was used to extract a unified video representation for all TTE views. The shared encoder enabled consistent feature learning while accommodating variable view availability across examinations. To handle missing views efficiently, only available views were processed via batch-wise concatenation, rather than by zero-padding absent inputs. The MViT-Small encoder, which incorporates multi-scale attention mechanisms, was initialized with weights pretrained on an in-house TTE view classification task to leverage domain-specific representations. For each input view, the encoder generated a fixed-length feature embedding, which was subsequently used for downstream MV etiology classification. Detailed input dimensions, batch-wise concatenation procedures, and view-specific feature separation are provided in **Supplementary Methods 3**.

To reflect established clinical interpretation workflows, encoder features from each view were processed through view-specific projection modules. Features derived from the PLAX view were decomposed into valve-specific and cavity-related representations, mirroring the clinical practice of separately assessing valvular morphology and ventricular context in this view. In contrast, the A4C and color Doppler views are typically interpreted by evaluating overall chamber geometry and flow dynamics as integrated information rather than isolating valve and cavity components. Accordingly, features from these views were projected into a shared representation space without further decomposition into valve and cavity components, allowing them to contribute complementary chamber-level and hemodynamic information that is less accessible from the PLAX perspective. PLAX-zoomed views were not directly used for classification; instead, they were incorporated to refine valve-specific representations through a learnable, zoom-aware gating mechanism. This mechanism dynamically integrated detailed zoomed morphological information when available, while defaulting to standard PLAX-derived valve features when zoomed views were absent. Additional methodological details are available in **Supplementary Methods 4.**

Final MV etiology classification was performed by integrating information from all available TTE views. The refined valve representation, cavity-related features from the PLAX view, and complementary features from the A4C and color Doppler views were concatenated to form a unified feature representation. When a view was unavailable, its corresponding feature position was filled with a learnable embedding. This embedding was used to explicitly represent the absence of that view during feature fusion, rather than to reconstruct or infer the missing image content. The fused representation was then passed to a linear classification head to predict one of five etiologic categories (normal, prolapse, rheumatic, functional, or degenerative mitral valve disease). The model was trained using a composite objective function that combined classification loss with auxiliary regularization terms to promote stable feature learning and effective use of zoomed morphological information. Detailed descriptions are provided in **Supplementary Method 5.**

The model was trained using the AdamW optimizer with learning rate scheduling and regularization strategies to promote stable convergence and prevent overfitting. The checkpoint achieving the best performance on the validation set was selected for final evaluation. Detailed training configurations are provided in **Supplementary Methods 6**.

### Validation and Statistical Analysis

Model performance was evaluated using standard classification metrics, including overall accuracy, balanced accuracy, and macro-F1 score as global performance metrics. In addition, class-specific diagnostic performance was assessed for each MV etiology using accuracy, precision, sensitivity, specificity, F1-score, and the area under the receiver operating characteristic curve (AUROC). Class-wise accuracy, precision, sensitivity, specificity, and F1-score were derived from confusion matrices using counts of true positives, true negatives, false positives, and false negatives. AUROC was calculated for each etiology using a one-vs-rest strategy, with receiver operating characteristic curves generated by varying classification thresholds based on predicted class probabilities. To evaluate the reliability of MV etiology labeling, an additional interobserver agreement analysis was performed using 60 TTE examinations sampled from the internal and external test datasets (30 each). Cases were selected so that the etiologic distribution of each sampled subset broadly reflected that of the corresponding test dataset. These examinations were independently reviewed by three cardiologists, and interobserver agreement for MV etiology classification was quantified using Fleiss’ kappa.^20^

To further investigate factors potentially contributing to performance differences between datasets, additional stratified analyses were performed in cases with at least mild MR. Specifically, model performance was evaluated separately in mild MR and moderate or greater MR subgroups. In addition, IQ-stratified analyses were also conducted to assess model robustness under variable imaging conditions. Automated IQ assessment was applied to B-mode TTE views (PLAX, PLAX zoomed MV, and A4C). Based on these assessments, examinations were categorized into an all-adequate group, in which all input views met at least fair IQ, and a partially suboptimal group, in which one or more input views were rated below the fair threshold. Model performance was compared between these groups using the same diagnostic metrics described above.

To qualitatively assess model interpretability, attention-based visualization was performed using self-attention weights extracted from the final layer of the MViT encoder. Attention maps were generated to identify spatiotemporal regions prioritized by the model during feature extraction. This approach was selected in preference to gradient-based visualization methods due to architectural constraints related to global pooling and multi-view feature fusion. Attention weights were averaged across heads and frames and overlaid on representative echocardiographic images to visualize anatomically relevant focus areas, such as valve leaflets and surrounding structures.

## RESULTS

### Baseline Characteristics of the Study Population

Baseline Characteristics and the distribution of MV pathology across datasets are shown in **Table 1**. Within the developmental dataset, the training, validation, and internal test subsets were well balanced with respect to age, sex, body mass index, prevalence of MS and MR, and MV etiology (all p >0.05). Compared with the internal test dataset, patients in the external test dataset were older and had a higher prevalence of both MS and MR. In addition, the distribution of MV etiologies differed significantly between the internal and external test datasets (p<0.001), with the external dataset showing a higher proportion of degenerative and functional etiologies and a lower proportion of prolapse. Furthermore, the severity distribution of MR differed between the developmental and external datasets, with the external cohort demonstrating a higher proportion of mild MR and a lower proportion of severe MR across multiple MV etiologies (**Figure 2**). In contrast, differences in MS severity distribution between datasets were less pronounced (**Supplementary Results 1**).

**Figure 2.**
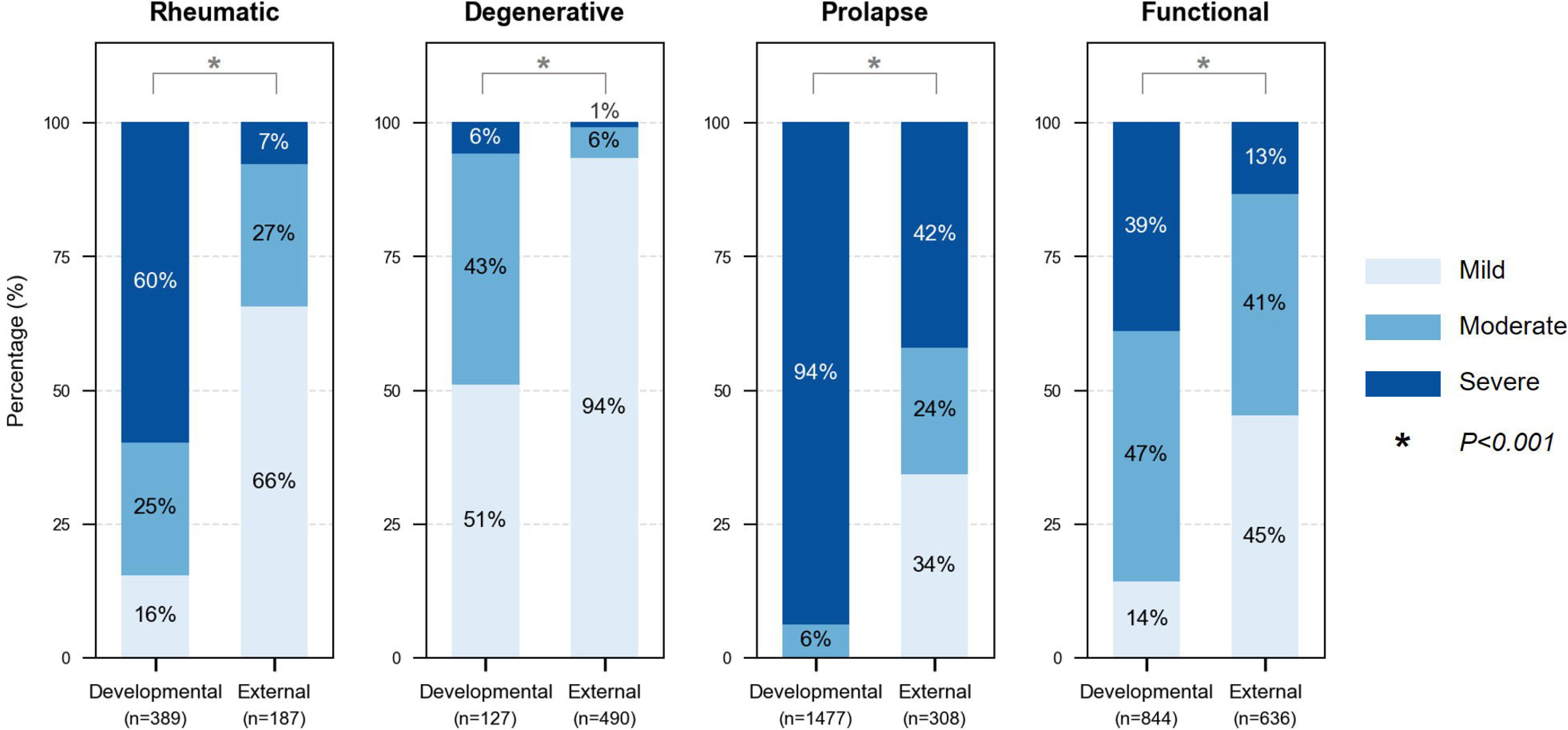
Mitral Regurgitation Severity Distribution in Developmental and External Cohorts. Distribution of mitral regurgitation (MR) severity (mild, moderate, and severe) across major etiologic categories in the developmental and external datasets. The external dataset showed a relative shift toward milder MR severity compared with the developmental dataset across several etiologies.

In an additional interobserver analysis using 60 TTE examinations sampled from the internal and external test datasets (30 each) with approximate preservation of their etiologic distributions, agreement among three cardiologists for MV etiology classification was high. Fleiss’ kappa values were 0.864 for the internal subset, 0.828 for the external subset, and 0.849 overall.

### Diagnostic Performance of the DL Model in Internal and External Datasets

In the internal test dataset (n=430), the model achieved an overall accuracy of 0.916, a balanced accuracy of 0.867, and a macro-F1 score of 0.857, indicating robust and well-balanced classification performance across etiological categories. In the external test dataset (n=2,262), overall performance remained high, with an overall accuracy of 0.813, a balanced accuracy of 0.792, and a macro-F1 score of 0.804, supporting the generalizability of the model across independent cohorts. The diagnostic performance of the proposed DL model for MV etiology classification is summarized using confusion matrices (**Figure 3)** and detailed quantitative performance metrics (**Table 2**). To facilitate the interpretation of class-specific error patterns, row-normalized confusion matrices are additionally provided in **Supplementary Results 2.**

**Figure 3.**
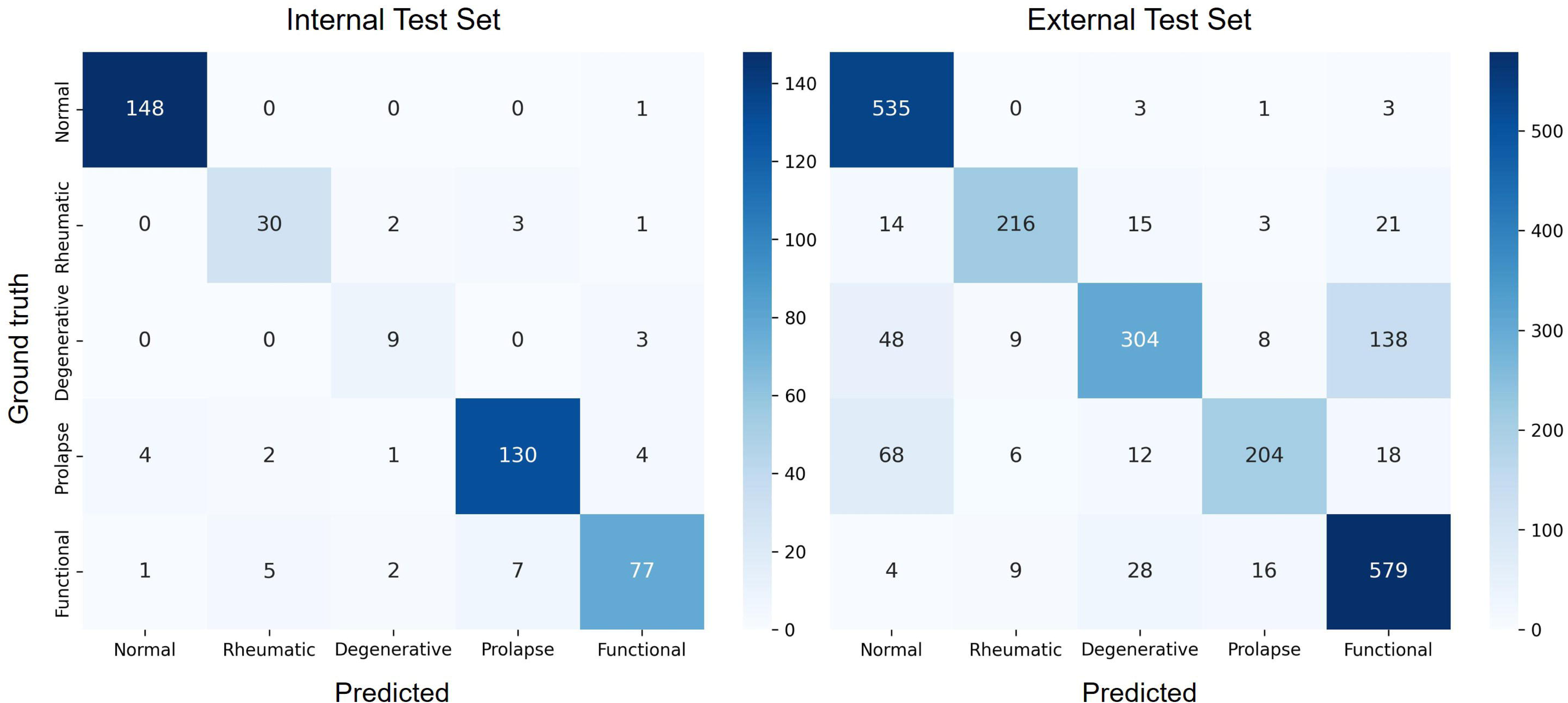
Confusion Matrices of Deep Learning-based Mitral Valve Etiology Classification in the Internal and External Test Datasets.

**Table 2.** Diagnostic Performance of the Proposed Deep Learning Model on Internal and External Test Datasets.

|  | Internal Test Dataset |  |  |  |  |  | External Test Dataset |  |  |  |  |  |
| --- | --- | --- | --- | --- | --- | --- | --- | --- | --- | --- | --- | --- |
|  | Accuracy | Precision | Sensitivity | Specificity | F1-score | AUROC | Accuracy | Precision | Sensitivity | Specificity | F1-score | AUROC |
| **Normal** | 0.986 | 0.967 | 0.993 | 0.982 | 0.980 | 0.997 | 0.938 | 0.800 | 0.987 | 0.922 | 0.884 | 0.992 |
| **Rheumatic** | 0.970 | 0.811 | 0.833 | 0.982 | 0.822 | 0.968 | 0.966 | 0.900 | 0.803 | 0.988 | 0.849 | 0.966 |
| **Degenerative** | 0.981 | 0.643 | 0.750 | 0.988 | 0.692 | 0.980 | 0.885 | 0.840 | 0.600 | 0.967 | 0.700 | 0.931 |
| **Prolapse** | 0.951 | 0.929 | 0.922 | 0.965 | 0.925 | 0.985 | 0.942 | 0.879 | 0.662 | 0.986 | 0.756 | 0.942 |
| **Functional** | 0.944 | 0.895 | 0.837 | 0.973 | 0.865 | 0.969 | 0.895 | 0.763 | 0.910 | 0.889 | 0.830 | 0.957 |

AUROC = area under the receiver operating characteristic curve

In the internal test dataset, excellent performance for normal valves was observed, with an accuracy of 0.986, a sensitivity of 0.993, and an AUROC of 0.997. High diagnostic performance was also observed for rheumatic disease (accuracy 0.970, AUROC 0.968) and degenerative disease (accuracy 0.981, AUROC 0.980), although degenerative cases showed relatively lower sensitivity compared with other etiologies. For prolapse, the model demonstrated high accuracy (0.951) and AUROC (0.985), with balanced precision and sensitivity. Functional MV disease was classified with an accuracy of 0.944 and an AUROC of 0.969, showing slightly lower sensitivity compared with normal valves but preserved overall discriminative performance.

In the external test dataset, overall diagnostic performance remained high, supporting the model’s generalizability across independent cohorts. Classification of normal valves remained robust, with an accuracy of 0.938 and an AUROC of 0.992. For rheumatic disease, the model maintained strong performance, achieving an accuracy of 0.966 and an AUROC of 0.966. Degenerative MV disease demonstrated slightly lower accuracy (0.885) but preserved discriminative ability (AUROC = 0.931), although sensitivity was reduced compared with other etiologies (0.600). Prolapse also showed a modest decline in sensitivity (0.662) in the external dataset, with an accuracy of 0.942 and an AUROC of 0.942. Functional MV disease demonstrated high sensitivity (0.910) and an AUROC of 0.957, indicating robust discriminative performance despite modestly reduced accuracy (0.895), comparable to that observed for degenerative disease. Confusion matrix analysis revealed that most misclassifications occurred between prolapse, degenerative, and functional etiologies, reflecting overlapping morphological and hemodynamic features among these categories, whereas normal and rheumatic etiologies were more consistently classified across both datasets. Additional exploratory UMAP-based visualization of encoder and fused feature representations in the external test dataset is provided in **Supplementary Results 3.**

Given the observed differences in diagnostic performance between the internal and external test datasets, additional analyses were performed after restricting the cohort to cases with at least mild MR and stratifying the remaining cases by MR severity. These severity-restricted analyses represent a more challenging classification setting than the full cohort because normal or trivial MR cases are excluded and the remaining cases are enriched for etiologies with substantial morphological overlap. In the internal test dataset, severity-stratified patterns were less uniform across etiologies, likely reflecting smaller subgroup sizes and dataset-specific mix. In the external dataset, overall model performance was higher in the subgroup with MR of moderate or greater severity than in the mild MR subgroup, with a particularly improved sensitivity for prolapse. Detailed results are presented in **Supplementary Results 4**.

Additional exploratory analyses were also performed according to MS severity. Because clinically significant MS is largely dominated by rheumatic and non-rheumatic degenerative etiologies, these analyses were interpreted primarily within that clinical framework and are presented in **Supplementary Results 5.**

To further clarify the contribution of color Doppler inputs, an additional supplementary analysis was performed using a B-mode–only model. Etiologic classification remained feasible using B-mode views alone, with broadly preserved performance in the internal test dataset and modest attenuation in the external test dataset, particularly among non-rheumatic MR etiologies. These findings suggest that B-mode views contain meaningful morphologic information for MV etiology classification, while color Doppler provides complementary information within an integrated routine echocardiographic assessment framework. Detailed results are provided in **Supplementary Results 6.**

### Performance According to View Completeness

To further assess robustness to incomplete input availability, we analyzed the distribution of missing input views and compared diagnostic performance between examinations with complete view and those with one or more missing views. Complete views were available in 266 of 430 internal examinations (61.9%) and 2,065 of 2,262 external examinations (91.3%). No clinically meaningful degradation in diagnostic performance was observed in incomplete-view studies in either cohort. Detailed missing-view distributions, combinations, and class-specific performance according to view completeness are provided in **Supplementary Results 7**.

### Performance in Cases with Multiple MV Pathologies

Although cases with multiple coexisting MV disease etiologies were excluded from the developmental dataset because a single dominant etiology could not be clearly determined, we performed a post-hoc inference analysis to examine model behavior in this clinically relevant subgroup. A total of 63 such cases were identified and analyzed. The most common mixed-etiology pattern was combined functional and degenerative MV disease, accounting for 18 of 63 cases; the detailed distribution of mixed etiologies is provided in **Supplementary Results 8**. Among all mixed-etiology cases, the DL model correctly predicted the expert-designated primary etiology in 42 cases (66.7%). In an additional 12 cases (19.0%), the model prediction corresponded to the secondary (comorbid) etiology identified by expert reviewers. Overall, the model predicted one of the expert-recognized etiologies in 54 of 63 cases (85.7%), despite not being trained using mixed-etiology labels. (**Supplementary Results 8**) The remaining 9 cases were misclassified into etiologic categories not identified by expert review. These cases typically demonstrate complex or overlapping morphological and hemodynamic features, highlighting the inherent ambiguity in assigning a single dominant etiology in advanced or mixed MV disease. Representative examples of model predictions in mixed-etiology cases are illustrated in **Figure 4**, with the corresponding cine loops provided in **Videos 1-4**.

**Figure 4.**
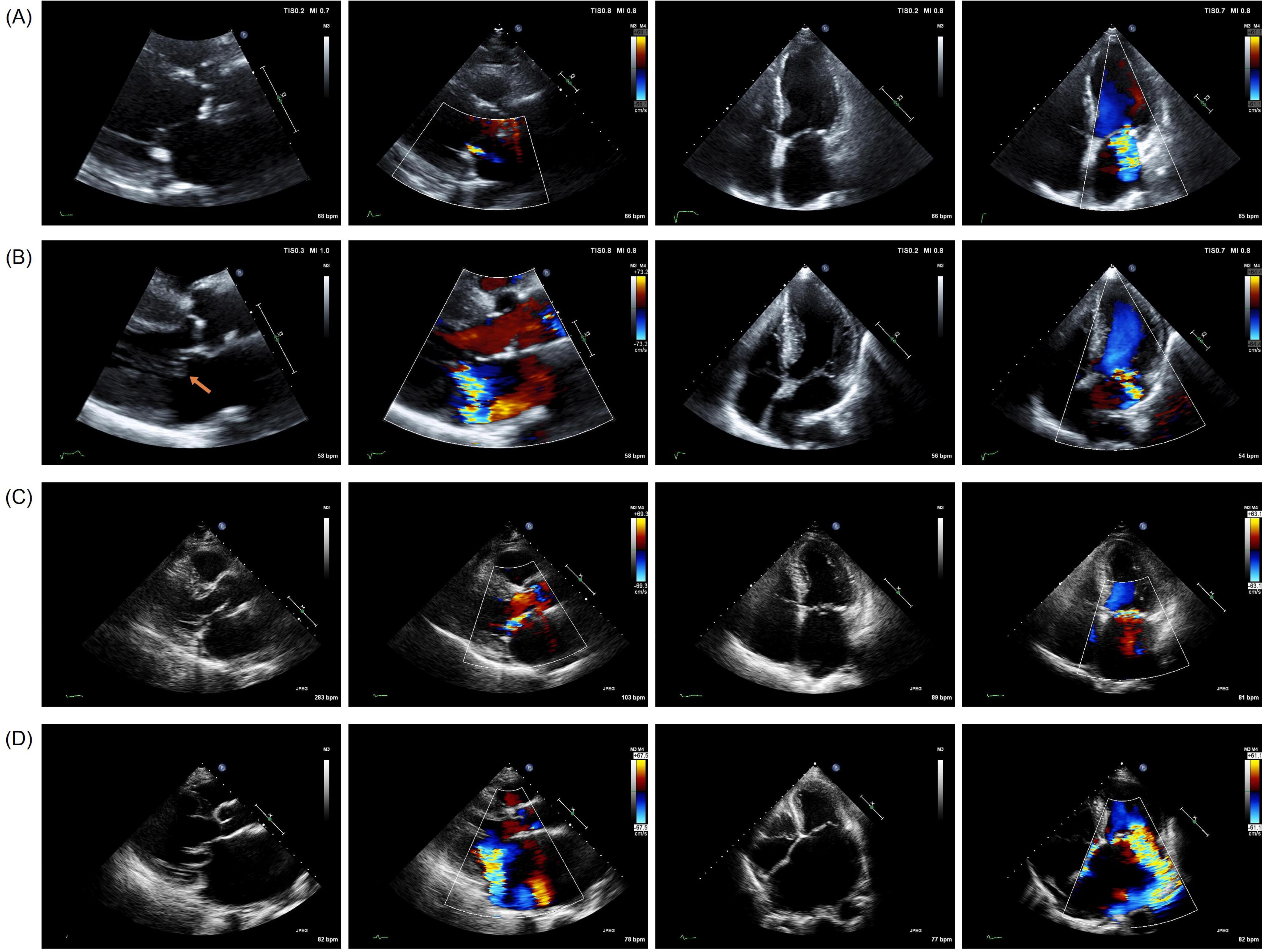
Representative Examples of Deep Learning-Based Predictions in Cases with Multiple Mitral Valve Etiologies. (A) A woman in her 90s with mixed functional and degenerative mitral regurgitation (MR). The patient had severe ischemic cardiomyopathy involving multivessel territories with a left ventricular ejection fraction (LVEF) of 29%, resulting in marked leaflet tethering consistent with functional MR. Concomitant mitral annular calcification (MAC) was also evident, suggesting degenerative changes. The deep learning (DL) model predicted functional mitral valve disease. (B) A man in his 80s with coexisting functional and prolapse mitral valve pathology. LVEF was preserved (66%); however, a regional wall motion abnormality of the mid-posterolateral wall was present, contributing to functional MR. In addition, prolapse of the anterior mitral leaflet was observed on the parasternal long-axis view. The DL model predicted mitral valve prolapse. (C) A man in his 60s with mixed prolapse and degenerative MR. Echocardiography showed severe eccentric MR caused by chordal rupture and flail motion of the middle scallop of the posterior mitral leaflet (P2), consistent with prolapse. In addition, marked MAC was present, supporting a coexisting degenerative component. LVEF was preserved (65%). The DL model predicted MV prolapse. (D) A man in his 70s with coexisting prolapse and functional MR. Echocardiography demonstrated flail motion of the medial portions of both mitral leaflets (A3 and P3) with chordal rupture, consistent with prolapse. At the same time, long-standing severe MR was associated with chronic left atrial enlargement and annular dilatation, contributing an additional functional component with generalized coaptation failure. LVEF was preserved (59%). The DL model predicted mitral valve prolapse. Corresponding videos for panels A–D are provided in Videos 1–4, respectively.

### Subgroup Analyses According to Image Quality

Model performance was further evaluated using IQ to assess robustness under varying image conditions. When examinations were stratified into all-adequate and partially suboptimal groups, overall diagnostic performance remained largely preserved across both internal and external test datasets. (**Supplementary Results 9**) In the internal test dataset, overall accuracy, balanced accuracy, and macro-F1 score were comparable between the two IQ subgroups (0.932 vs. 0.883; 0.853 vs. 0.879; and 0.849 vs. 0.859, respectively), with no systematic degradation in class-wise performance observed in the partially suboptimal group. Consistent findings were observed in the external test dataset. Overall accuracy, balanced accuracy, and macro-F1 score were similar between the all-adequate and partially suboptimal groups (0.808 vs. 0.825; 0.776 vs. 0.829; and 0.791 vs. 0.823, respectively), indicating that incomplete or partially suboptimal IQ did not result in a clinically meaningful decline in model performance. Across etiologic categories, sensitivity and AUROC values remained broadly comparable across image quality strata, supporting the robustness of the proposed model in real-world imaging conditions in which optimal view acquisition may not always be feasible.

### Attention Map-Based Model Interpretability Analysis

Attention map analysis was conducted to qualitatively assess whether the model focused on anatomically relevant regions during feature extraction. Representative cases containing all five target views were selected for each MV etiology category. As shown in **Figure 5**, the model predominantly attended to the MV apparatus across all categories and views, including normal valves. In PLAX and PLAX-zoomed views, attention was predominantly concentrated on the MV leaflet and annular regions. In A4C views, attention remained largely focused on the MV apparatus. In color Doppler views, attention extended beyond valvular anatomy to include regions corresponding to regurgitant flow in cases with MR. These attention patterns were consistently observed across prolapse, rheumatic, functional, and degenerative MV etiologies. Overall, the attention maps suggest that the model prioritizes anatomically and physiologically relevant regions when extracting features for MV etiology classification.

**Figure 5.**
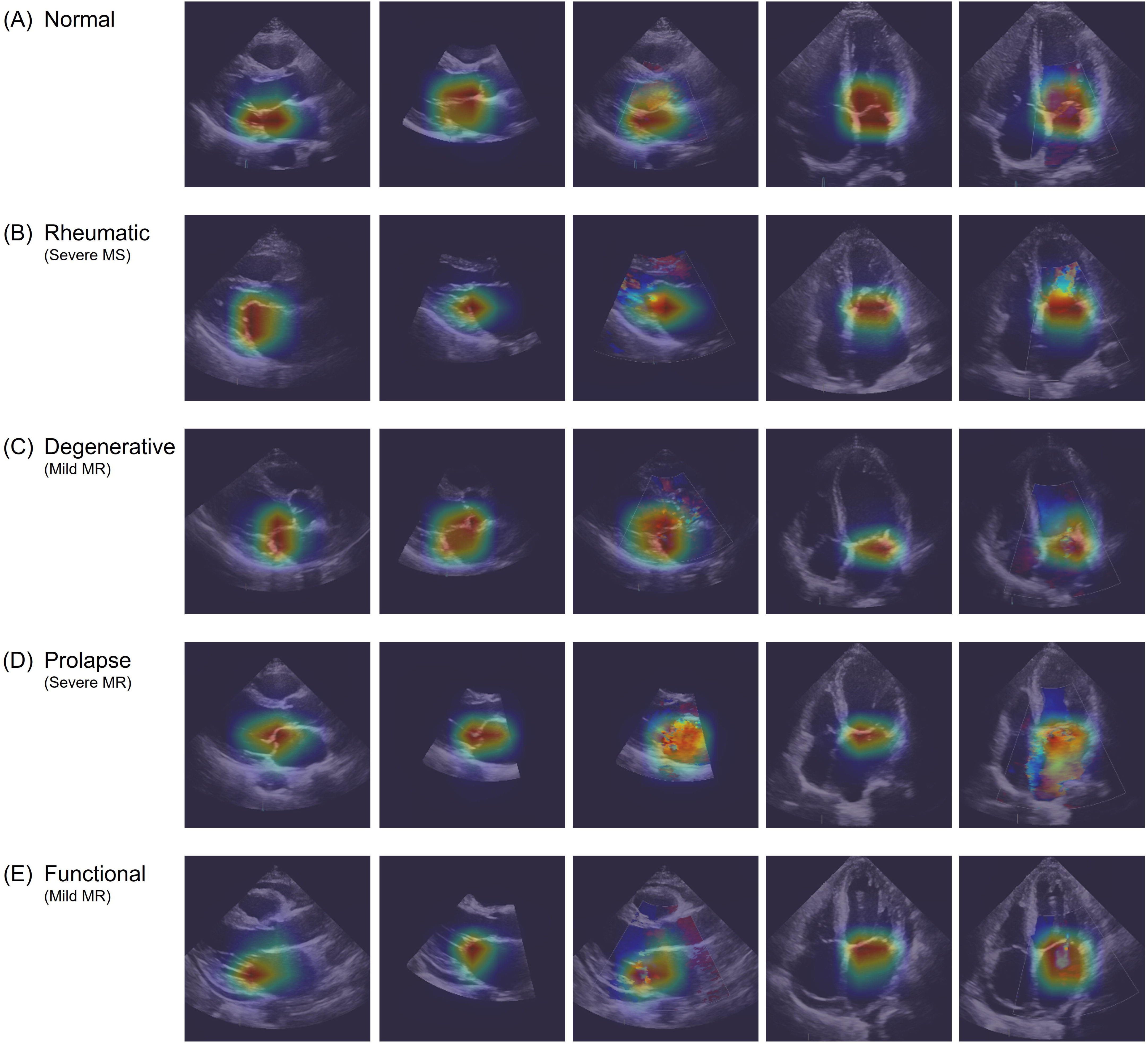
Attention Maps Highlighting Anatomically Relevant Regions for Mitral Valve Etiology Classification. Attention maps illustrate regions predominantly weighted by the model across routine echocardiographic views during mitral valve etiology inference.

## DISCUSSION

In this study, we developed a DL-based framework for the classification of normal and four major MV disease etiologies using a limited set of routinely acquired B-mode and color Doppler TTE views. The model demonstrated stable and reproducible performance in both internal and external test cohorts, supporting its generalizability across independent clinical settings. Diagnostic performance was particularly high for normal valves and rheumatic MV disease, while overall accuracy and discriminative ability were largely preserved in the external cohort despite differences in disease severity distribution. Importantly, model performance remained robust across analyses stratified by MR severity, IQ, and the presence of mixed MV pathologies. Together, these findings suggest the feasibility of inferring MV etiology from limited echocardiographic inputs under routine clinical imaging conditions.

AI-based echocardiographic analysis has rapidly evolved in recent years. Early applications primarily focused on automating quantitative measurements^5–8,21,22^, whereas more recent approaches have expanded toward replicating expert visual interpretation.^23,24^ These advances have been applied to VHD, initially automating diagnosis and severity grading based on quantitative parameters, most notably for aortic stenosis,^6,25,26^ and subsequently by directly analyzing echocardiographic videos to detect VHD and estimate severity, either through valve-specific algorithms^6,11,27–29^ or as part of a full-examination framework.^30,31^ More recently, Al-Alusi et al. demonstrated that DL can accurately identify MV prolapse directly from echocardiographic videos, further supporting the feasibility of AI-based morphologic interpretation of MV disease.^32^ However, automated multiclass etiologic classification across major MV disease categories remains challenging. This limitation is particularly relevant in MV disease, where accurate etiologic assessment requires integration of leaflet and subvalvular features and has direct implications for clinical management. In this context, we developed and validated a DL-based framework designed to infer MV etiology from B-mode and color Doppler inputs under routine clinical imaging conditions. Because echocardiographic etiologic assessment of MV disease in routine practice is typically based on integration of both structural and flow-related information, color Doppler views were intentionally included as complementary inputs rather than as substitutes for morphologic assessment. Accordingly, the present framework should be interpreted as reflecting integrated routine echocardiographic assessment rather than purely morphology-only classification, and the model may therefore have used hemodynamic correlates of valve pathology in addition to structural features.

In the present study, etiology-specific analyses provided insight into the observed performance patterns. Diagnostic separability was highest for normal valves and rheumatic MV disease, reflecting their relatively distinct morphological features. In contrast, performance varied among non-rheumatic MR etiologies and was differentially influenced by disease severity. Sensitivity for MV prolapse improved in cases with MR of moderate or greater severity, consistent with clearer expression of characteristic morphologic and hemodynamic features. By contrast, degenerative MV disease did not show comparable improvement, but instead demonstrated lower sensitivity in this subgroup. This pattern may reflect the intrinsic heterogeneity of degenerative MV disease and its substantial phenotypic overlap with prolapse and functional MR. In the external test cohort, performance attenuation among non-rheumatic MR etiologies may also reflect the real-world labeling framework, in which each case was assigned a single predominant etiology even when overlapping morphologic features were present. Post-hoc analyses of mixed-etiology cases further support a continuum among non-rheumatic MR etiologies, as the model frequently identified either the primary or secondary expert-assigned etiology. Collectively, these findings suggest that the DL model captures latent etiologic structure within echocardiographic morphology, while its performance is shaped by lesion severity, phenotypic overlap, and dataset-specific case mix.

The proposed framework demonstrated stable diagnostic performance across varying IQ conditions, with no clinically meaningful degradation observed in examinations with partially suboptimal views. Notably, the slightly higher global metrics observed in the partially suboptimal subgroup should not be interpreted as evidence that lower IQ improves model performance. Rather, this finding likely reflects the composition of the subgroup, which included not only rare IQ-low examinations, defined as studies in which all available views were classified as poor or non-diagnostic, but also mixed-quality examinations in which fair-or-better views coexisted with poorer quality views. This robustness likely reflects the model design, which explicitly accommodates incomplete or lower-quality inputs through multi-view feature integration and learnable representations for missing views. Importantly, IQ was assessed using an automated, model-based approach rather than subjective visual grading, enhancing objectivity and reproducibility. These findings are clinically relevant, as real-world TTE acquisition is frequently constrained by patient-related factors and variable examination conditions. The preserved performance under suboptimal IQ supports the feasibility of applying the proposed model in routine clinical workflows, where optimal visualization of all target views cannot be consistently guaranteed.

The ability to infer MV etiology from limited, routinely acquired TTE views has several potential clinical implications. In routine, accurate etiologic assessment of MV disease often depends on operator expertise and may be challenging outside specialized centers. An AI-based framework capable of providing consistent etiologic suggestions could support early diagnostic triage and structured reporting, particularly in high-volume or resource-limited settings. By differentiating among major MV disease etiologies, the proposed model may help identify cases that warrant further expert review or advanced imaging. In addition, automated etiologic classification could complement existing AI tools focused on quantitative measurement and severity grading, contributing to more standardized and reproducible MV assessment. Collectively, these findings suggest that DL-based etiologic analysis may serve as a supportive component within AI-augmented echocardiographic workflows. At the same time, its clinical use requires caution. In particular, less experienced users may be vulnerable to over-reliance on AI-generated suggestions without sufficient critical appraisal, potentially weakening the gradual learning process needed to develop echocardiographic expertise. Therefore, such tools should be implemented in a manner that preserves clinician oversight and encourages integration of AI outputs with comprehensive image interpretation and clinical context.

Several limitations of this study should be acknowledged. First, the proposed framework was developed under the assumption of a single dominant MV etiology, whereas mixed or overlapping pathologies are common in clinical practice. Although post-hoc analyses suggested that the model captures overlapping etiologic features, performance in such cases remains inherently constrained by single-label training and evaluation. Second, etiologic classification was performed using a limited set of standard echocardiographic views rather than the full transthoracic echocardiographic examination. While this design choice reflects routine clinical acquisition and enhances feasibility, a comprehensive etiologic assessment may benefit from integrating additional views and complementary quantitative information. In particular, although zoomed A4C views focused on the MV may provide useful complementary morphologic information, they were not included because the current automatic view identification algorithm does not yet reliably distinguish this view category.^5–8,18^ In addition, because color Doppler views were included to reflect integrated routine echocardiographic interpretation, the model may have relied in part on hemodynamic correlates of valve pathology in addition to purely structural morphology. Building on experience with the current framework, we are developing an advanced model to analyze the full echocardiographic examination by integrating vision-based interpretation and quantitative measurements. Third, to prevent patient-level data leakage, repeated TTE examinations from the same patient were assigned exclusively to the training set. Although this design may have introduced some enrichment of follow-up cases in the training dataset, the developmental cohort was derived from a multicenter curated database designed to cover normal studies and a broad spectrum of cardiovascular disease rather than a consecutive institutional cohort, and repeated examinations were limited in number, involving only 23 patients and 50 TTE examinations among 3,485 TTE examinations. Therefore, the resulting selection bias was likely modest. Fourth, because this study represented an initial attempt at morphology-based deep learning classification of MV etiology, we applied conservative exclusion criteria to minimize potential imaging confounders related to prior surgery. As a result, cases with postoperative or peri-mitral structural alterations, such as prior tricuspid annuloplasty or ascending aorta replacement, were excluded, and the performance of the model in these settings remains to be established. Fifth, external validation relied on real-world clinical labels assigning a single predominant etiology, which were reviewed and confirmed by a single cardiologist rather than through multi-reader adjudication. This design better reflected routine clinical practice, but may have contributed to label uncertainty and performance attenuation for non-rheumatic MR etiologies characterized by substantial morphological overlap. Sixth, although the proposed framework demonstrated promising diagnostic performance, the present study was not designed to establish its clinical utility or incremental value over expert echocardiographic interpretation. Future prospective studies are needed to directly compare model performance with expert readers and to determine whether this approach improves diagnostic consistency, workflow efficiency, or downstream clinical decision-making. Finally, although attention-based visualization provided qualitative insight into model behavior, it does not establish causal relationships between specific imaging features and model predictions and should be interpreted as an explanatory aid rather than definitive evidence of feature importance.

## CONCLUSIONS

In this study, we demonstrated that DL–based analysis of limited, routinely acquired B-mode and color Doppler TTE views can support etiologic classification of MV disease. The proposed framework achieved stable performance across internal and external cohorts and remained robust under variations in disease severity, image quality, and etiologic overlap. These findings support the feasibility of AI-based etiologic classification as a complement to quantitative automation and expert visual assessment. Further studies are needed to establish its clinical utility and incremental value in routine echocardiographic practice.

## Data Availability

The AI-Hub dataset used in this study may be available upon proper request and approval of a formal proposal. The external test dataset cannot be made publicly shared due to ethical restrictions imposed by the IRB of the study institution, as public access could compromise patient confidentiality and privacy. Researchers interested in accessing the minimal anonymized dataset may contact the corresponding author for further information.

https://aihub.or.kr

## Contributors

**Dawun Jeong:** Data Curation, Methodology, Formal Analysis

**Moon-Seung Soh:** Validation, Investigation, Data Curation, Echocardiographic review, and annotation

**Jaeik Jeon:** Software, Methodology, Resources **Hyunseok Jeong:** Data Curation, Methodology **JunHeum Cho:** Software, Methodology, Resources **Jiyeon Kim:** Software, Methodology, Resources **Jina Lee:** Data Curation, Methodology

**Seung-Ah Lee:** Investigation, Supervision, Echocardiographic review, and annotation

**Yeonggul Jang:** Software supervision, Technological methodology oversight

**Yeonyee E. Yoon:** Conceptualization, Supervision, Echocardiographic review and annotation, Clinical interpretation, Writing, Review, and Editing

**Hyuk-Jae Chang:** Conceptualization, Supervision, Funding acquisition All authors read and approved the final version of the manuscript.

## Acknowledgments

This work was supported by a grant from the Institute of Information & Communications Technology Planning & Evaluation (IITP) funded by the Korean government (Ministry of Science and ICT) (No.2022000972, Development of a Flexible Mobile Healthcare Software Platform Using 5G MEC); and the Medical AI Clinic Program through the NIPA funded by the MSIT. (Grant No.: H0904-24-1002).

## Data Sharing Statement

The AI-Hub dataset used in this study may be available upon proper request and approval of a formal proposal. The external test dataset cannot be publicly shared due to ethical restrictions imposed by the IRB of the study institution, as public access could compromise patient confidentiality and privacy. Researchers interested in accessing the minimal anonymized dataset may contact the corresponding author for further information.

---

**Central Illustration.**
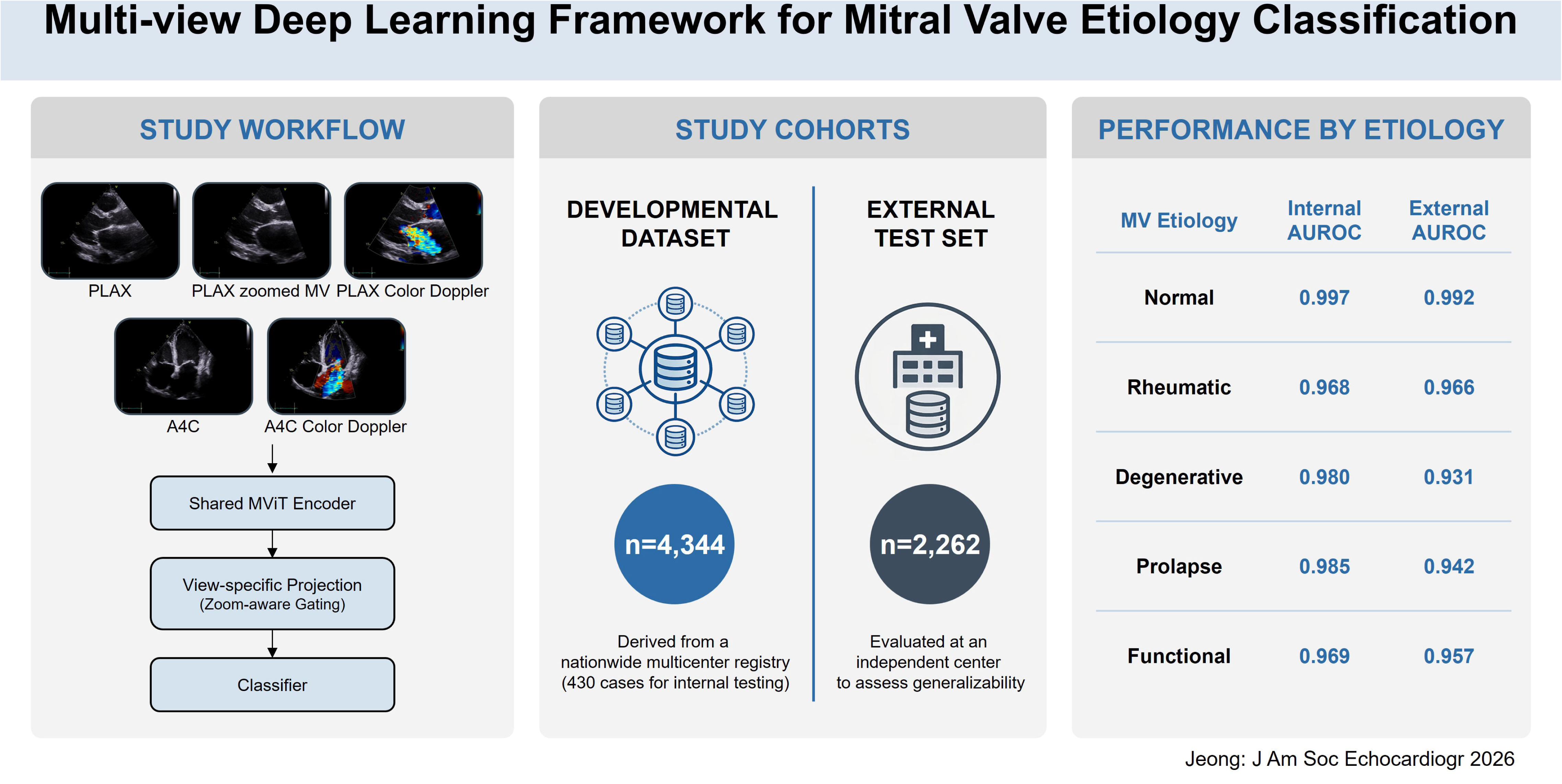
Multi-view Deep Learning Framework for Mitral Valve Etiology Classification. The proposed deep learning framework integrates routinely acquired transthoracic echocardiographic views, including parasternal long-axis, zoomed parasternal long-axis mitral valve, parasternal long-axis color Doppler, apical 4-chamber, and apical 4-chamber color Doppler images, to classify mitral valve etiology. A shared MViT encoder first extracts common image features, followed by view-specific projection with zoom-aware gating and final etiologic classification. The model was developed using 4,344 cases from a nationwide multicenter registry and externally tested in 2,262 cases from an independent center. The framework demonstrated consistently high performance across major mitral valve etiologies, including normal, rheumatic, degenerative, prolapse, and functional mitral valve disease. Abbreviations: A4C, apical 4-chamber; AUROC, area under the receiver operating characteristic curve; MV, mitral valve; PLAX, parasternal long-axis.

## VIDEO LEGENDS

**Video 1.** Representative echocardiographic cine loops corresponding to Figure 4A, showing mixed functional and degenerative MR with leaflet tethering and concomitant mitral annular calcification.

**Video 2.** Representative echocardiographic cine loops corresponding to Figure 4B, showing coexisting functional MR and anterior mitral leaflet prolapse.

**Video 3.** Representative echocardiographic cine loops corresponding to Figure 4C, showing mixed prolapse and degenerative MR with P2 flail motion and marked mitral annular calcification.

**Video 4.** Representative echocardiographic cine loops corresponding to Figure 4D, showing coexisting prolapse and functional MR with flail motion of A3 and P3 and annular dilatation-related coaptation failure.

