## supplementary for "Deep Learning–Based Multiclass Classification of Mitral Valve Etiologies Using Limited B-Mode and Color Doppler Echocardiography: Internal and External Validation"


| Material | Page number |
|---|---|
| **Supplementary Methods 1. View Classification Module and Target Views Used in This Study** | 3 |
| **Supplementary Methods 2. Segmentation-Based Image Quality Assessment** | 6 |
| **Supplementary Methods 3. Shared Spatiotemporal Encoder and Batch-wise View Processing** | 9 |
| **Supplementary Methods 4. View-Specific Feature Projection and Zoomed-Aware Refinement** | 11 |
| **Supplementary Methods 5. Feature Fusion, Classification, and Training Objective** | 13 |
| **Supplementary Methods 6. Training Strategy** | 15 |
| **Supplementary Results 1. Mitral Stenosis Severity Distribution in the Developmental and External Datasets** | 16 |
| **Supplementary Results 2. Row-Normalized Confusion Matrices for MV Etiology Classification in the Internal and External Test Datasets.** | 17 |
| **Supplementary Results 3. UMAP-Based Characterization of Learned Feature Representations in the External Test Dataset** | 18 |
| **Supplementary Results 4. MR Severity-Stratified Diagnostic Performance of the DL Model in the Internal and External Test Dataset** | 20 |

| | |
|---|---|
| **Supplementary Results 5. MS Severity-Stratified Diagnostic Performance of the DL Model in the Internal and External Test Datasets** | 24 |
| **Supplementary Results 6. Diagnostic Performance of the B-Mode–Only Model in the Internal and External Test Datasets** | 27 |
| **Supplementary Results 7. Distribution of Missing Input Views and Performance According to View Completeness** | 29 |
| **Supplementary Results 8. Distribution and Model Performance in Cases with Multiple Mitral Valve Etiologies** | 34 |
| **Supplementary Results 9. Image Quality-Stratified Performance Analysis.** | 36 |

**Supplementary Methods 1. View Classification Module and Target Views Used in This Study**

The in-house echocardiographic view classification module was originally developed to recognize a broad range of echocardiographic views across multiple imaging modes, including B-mode/Color Doppler, M-mode, and spectral/tissue Doppler. In the present study, however, only the B-mode and Color Doppler view categories relevant to the mitral valve etiology classification framework were used. Automated view selection was first performed by this module, after which investigators reviewed the selected clips to remove misclassified views and recover missing target views. The target view categories and internal test performance of the B-mode/Color Doppler view classifier used in this study are summarized below.

**1.1.Target View Families and Imaging Modalities Used in the View Classification Module**

| **Echocardiographic views** | **B-mode** | **Color Doppler** |
| --- | --- | --- |
| Parasternal long-axis left ventricle | ✓ | ✓ |
| Parasternal long-axis zoomed AV | ✓ | ✓ |
| Parasternal long-axis zoomed AV & MV | ✓ | ✓ |
| Parasternal long-axis zoomed MV | ✓ | ✓ |
| Parasternal long-axis zoomed Aorta | ✓ | ✓ |
| Parasternal short-axis, level of great vessels | ✓ | ✓ |
| Parasternal short-axis, level of mitral valve | ✓ | ✓ |
| Parasternal short-axis, level of papillary muscle | ✓ | ✓ |
| Parasternal short-axis, level of apex | ✓ | ✓ |
| Apical four-chamber | ✓ | ✓ |
| Apical four-chamber zoomed left ventricle | ✓ | ✓ |
| Apical four-chamber right ventricular-focused | ✓ | ✓ |
| Apical five-chamber | ✓ | ✓ |
| Apical five-chamber zoomed left ventricle | ✓ | ✓ |
| Apical two-chamber | ✓ | ✓ |
| Apical two-chamber zoomed left ventricle | ✓ | ✓ |
| Apical three-chamber | ✓ | ✓ |

| | | |
|---|---|---|
| Apical three-chamber zoomed left ventricle | ✓ | ✓ |
| Subcostal four-chamber | ✓ | ✓ |
| Subcostal long-axis IVC | ✓ | ✓ |

In the present MV etiology classification framework, PLAX-zoomed views containing the MV apparatus were used as target inputs; therefore, both “PLAX zoomed MV” and “PLAX zoomed AV & MV” categories were eligible for the PLAX-zoomed MV-related input.

### 1.2.Performance in view classification

| Echocardiographic views | n | Precision | Recall | F1-score |
|---|---|---|---|---|
| Parasternal long-axis left ventricle | 768 | 0.990 | 0.999 | 0.994 |
| Parasternal long-axis zoomed AV & MV | 393 | 0.953 | 0.987 | 0.970 |
| Parasternal long-axis zoomed MV | 217 | 0.952 | 0.917 | 0.934 |
| Parasternal long-axis left ventricle (color) | 672 | 0.999 | 1.000 | 0.999 |
| Apical four-chamber | 631 | 0.994 | 0.995 | 0.994 |
| Apical four-chamber (color) | 802 | 0.985 | 0.993 | 0.989 |
| Other | 15,220 | 0.999 | 0.998 | 0.999 |

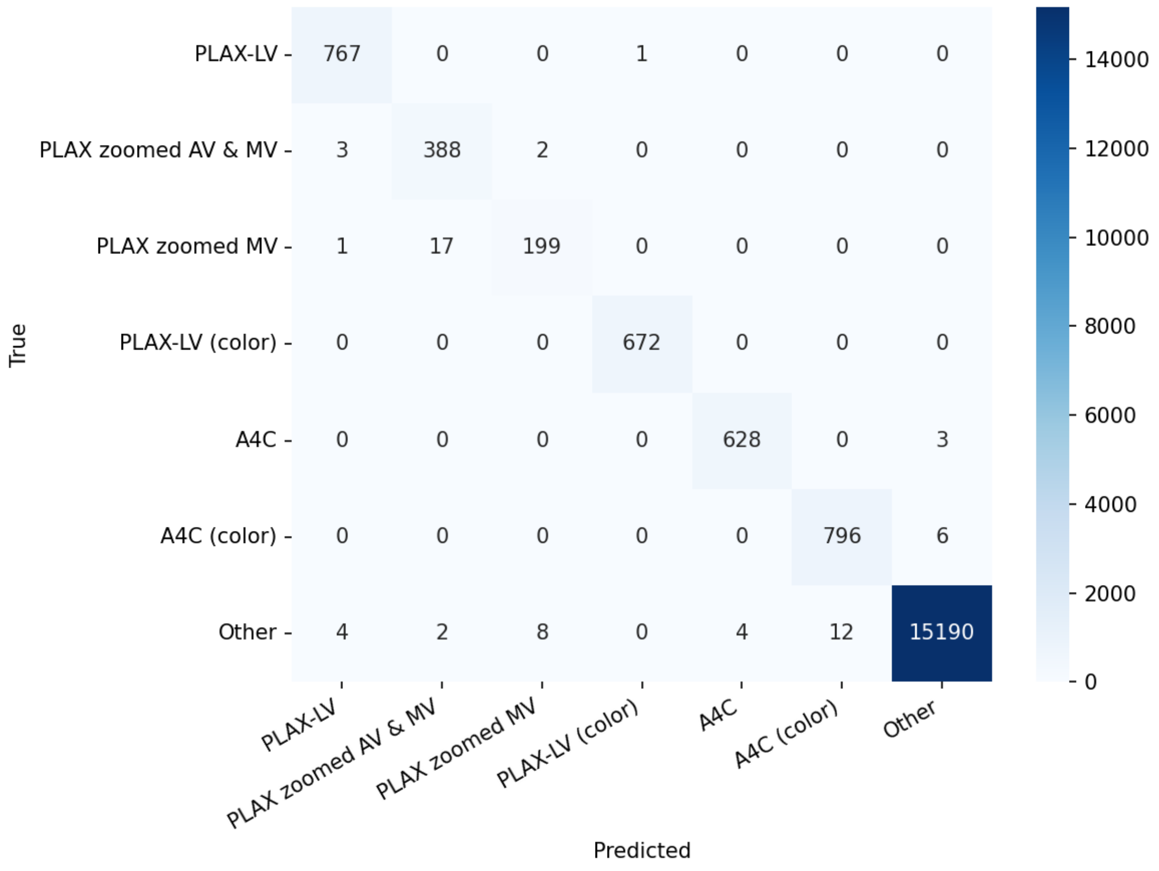

True
Predicted
PLAX-LV
PLAX zoomed AV & MV
PLAX zoomed MV
PLAX-LV (color)
A4C
A4C (color)
Other
767 0 0 1 0 0 0
3 388 2 0 0 0 0
1 17 199 0 0 0 0
0 0 0 672 0 0 0
0 0 0 0 628 0 3
0 0 0 0 0 796 6
4 2 8 0 4 12 15190
14000
12000
10000
8000
6000
4000
2000
0

### Supplementary Methods 2. Segmentation-Based Image Quality Assessment

Image quality (IQ) of transthoracic echocardiography (TTE) video clips was quantified using segmentation uncertainty derived from a deep learning (DL)–based anatomical segmentation model.[1-5] The model was originally developed to operate on standard B-mode TTE views, including parasternal long-axis view (PLAX), apical four-chamber view (A4C), apical two-chamber view (A2C), and apical three-chamber view (A3C), and generated pixel-wise class probability maps for each frame. For a clip consisting of $T$ frames with spatial resolution $H \times W$, the model outputs pixel-wise class probability $P \in [0, 1]^{T \times H \times W \times C}$ where C denotes the number of anatomical classes and $\sum_{c=1}^{C} p_{t,h,w,c} = 1$.

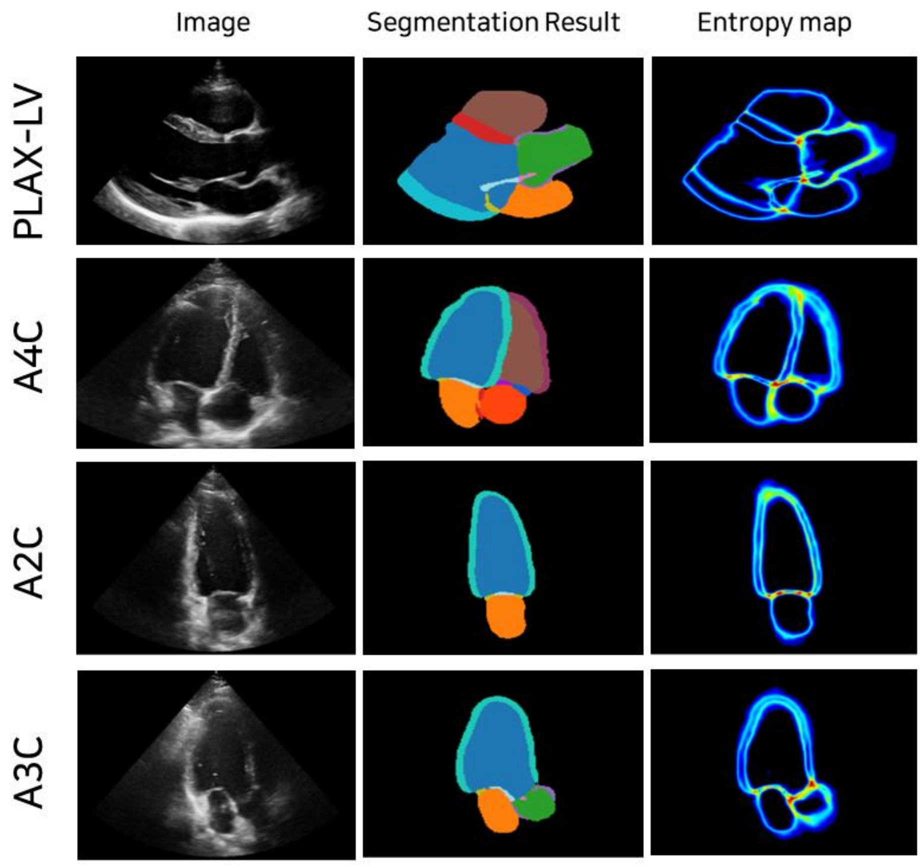


**Representative examples of the input image, predicted segmentation, and the corresponding entropy.**

To quantify per-pixel uncertainty, a Shannon entropy map was computed for each frame:

$$H_{t,h,w} = -\sum_{c=1}^{C} p_{t,h,w,c} \log\left(p_{t,h,w,c} + \epsilon\right)$$

where $\epsilon$ is a small constant added for numerical stability. Clip-level uncertainty was summarized as the mean entropy

$$\bar{H} = \frac{1}{|\Omega|} \sum_{(t,h,w)\in\Omega} H_{t,h,w}$$

calculated over the evaluated pixels set $\Omega$, defined as the minimal axis-aligned bounding box covering the predicted foreground region. This restriction was applied to mitigate dilution of entropy value by background regions. Higher value of $\bar{H}$ indicates greater segmentation uncertainty, and consequently lower IQ.

For calibration of the IQ metric, we used 453 B-mode TTE video clips acquired from 100 adult patients. Each clip was independently rated by two experienced sonographers, both Registered Diagnostic Cardiac Sonographers (RDCS) into four ordinal IQ categories (Good, Fair, Poor and Non-diagnostic) based on overall diagnostic usability. Rating considered the visibility of key anatomical structures (e.g., left ventricle, mitral valve, and apex), the presence of artifacts, and overall interpretability, following American Society of Echocardiography (ASE) recommendations.[6] Discrepancies between raters were resolved through discussion to achieve a consensus label, which served as the reference standard. The annotation process was supervised by board-certified cardiologists with more than 10 years of echocardiography experience (HJ Chang, MD, PhD; SA Lee, MD, PhD).

Using these expert labels, view-specific entropy thresholds $\tau_v^{(1)} < \tau_v^{(2)} < \tau_v^{(3)}$ were determined to map the continuous entropy-derived score to the four IQ categories as follows:

$$\hat{y}_v = \begin{cases} good, & \bar{H} < \tau_v^{(1)} \\ fair, & \tau_v^{(1)} \le \bar{H} < \tau_v^{(2)} \\ poor, & \tau_v^{(2)} \le \bar{H} < \tau_v^{(3)} \\ non-diagnostic, & \bar{H} \ge \tau_v^{(3)} \end{cases}$$

Within the same calibration cohort, the discriminative ability of the continuous entropy-based image quality score—rather than its external generalization performance—was evaluated by computing binary areas under the receiver operating characteristic curve (AUROC) at three ordinal decision boundaries: (1) non-diagnostic versus higher image quality, (2) poor or worse versus fair or better, and (3) fair or worse versus good. The boundary-specific AUROC values (in the same order) were as follows: PLAX, 0.744 / 0.749 / 0.918; A4C, 0.705 / 0.743 / 0.981; A2C, 0.765 / 0.775 / 0.999; and A3C, 0.701 / 0.708 / 0.993.

**Supplementary Methods 3. Shared Spatiotemporal Encoder and Batch-wise View Processing**

A shared spatiotemporal encoder based on the MViT-Small[7] architecture was employed to extract spatiotemporal feature representations from multiple echocardiographic views. The encoder was designed to operate on short video clips and to generate a fixed-length embedding for each input view, enabling consistent representation learning across heterogeneous echocardiographic inputs. Each input video clip was resized to a spatial resolution of $224 \times 224$ pixels and temporally sampled to 16 consecutive frames. Clips were represented as tensors of dimension

$$x \in \mathbb{R}^{B\times3\times16\times224\times224},$$

where $B$ denotes the batch size, and 3 represents RGB channels (applied uniformly to both grayscale B-mode and color Doppler images). The shared encoder mapped each input clip to a 512-dimensional feature embedding.

Because the availability of echocardiographic views varied across examinations, a batch-wise concatenation strategy was adopted to handle missing views without introducing artificial signals. Specifically, only available views were forwarded through the shared encoder, while absent views were excluded from processing rather than being represented by zero-padded inputs. Let $B$ denote the number of PLAX views available for all examinations, and let $n_z$, $n_a$, $n_{cp}$, and $n_{ca}$ denote the number of available PLAX-zoomed, apical four-chamber (A4C), PLAX color Doppler, and A4C color Doppler views, respectively. The concatenated input tensor was defined as

$$x_{\text{concat}} \in \mathbb{R}^{(B+n_z+n_a+n_{cp}+n_{ca})\times3\times16\times224\times224},$$

which was processed in a single forward pass through the shared encoder:

$$z_{\text{concat}} = f_{\theta_e}(x_{\text{concat}}) \in \mathbb{R}^{(B+n_z+n_a+n_{cp}+n_{ca})\times 512},$$

where $f_{\theta_e}(\cdot)$ denotes the shared encoder parameterized by $\theta_e$.

Following encoding, the resulting feature embeddings were separated back into view-specific tensors corresponding to each echocardiographic view. PLAX embeddings were available for all $B$ examinations ($B \times 512$), whereas PLAX-zoomed, A4C, PLAX color Doppler, and A4C color Doppler embeddings were retained only for examinations in which those views were present ($n_z \times 512$, $n_a \times 512$, $n_{cp} \times 512$, and $n_{ca} \times 512$, respectively). Binary masks were used to track view availability and to align view-specific embeddings with their corresponding examinations. This batch-wise processing strategy ensured that missing views did not interfere with representation learning while minimizing redundant computation compared with zero-padding–based approaches, which would otherwise introduce uninformative inputs.

The shared encoder was initialized using weights pretrained on an in-house echocardiographic view classification task, enabling transfer of domain-specific spatiotemporal representations. During training of the MV etiology classification framework, the encoder parameters were fine-tuned jointly with downstream network components.

**Supplementary Methods 4. View-Specific Feature Projection and Zoomed-Aware Refinement**

To enable anatomical specialization across echocardiographic views, encoder outputs were transformed using view-specific projection heads. Each projection head consisted of a two-layer multilayer perceptron (MLP) with Layer Normalization and ReLU activation, defined as:

$$g(h) = \mathrm{Linear}_{256\to256}\left(\mathrm{ReLU}\left(\mathrm{LayerNorm}\big(\mathrm{Linear}_{512\to256}(h)\big)\right)\right)$$

where the input dimension is 512 and both the hidden and output dimensions are 256.

Encoder features from the PLAX view $\mathrm{h}_p$ were decomposed into valve and cavity embeddings:

$$\mathrm{v} = g_v(\mathrm{h}_p) \in \mathbb{R}^{B\times F}, \mathrm{c} = g_c(\mathrm{h}_p) \in \mathbb{R}^{B\times F}$$

For optional views, only available samples were projected:

$$\mathrm{z} = g_z(\mathrm{h}_z) \in \mathbb{R}^{n_z\times F}, \mathrm{a} = g_a(\mathrm{h}_a) \in \mathbb{R}^{n_a\times F},$$

$$cp = g_{cp}(\mathrm{h}_{cp}) \in \mathbb{R}^{n_{cp}\times F}, ca = g_{ca}(\mathrm{h}_{ca}) \in \mathbb{R}^{n_{ca}\times F}$$

where $\mathrm{h}_z$, $\mathrm{h}_a$, $\mathrm{h}_{cp}$, and $\mathrm{h}_{ca}$denote encoder features from PLAX-zoomed, A4C, PLAX color Doppler, and A4C color Doppler views, respectively.

To robustly handle missing views at the feature level, each view maintained a learnable missing embedding $\mathrm{e}_{\mathrm{missing}} \in \mathbb{R}^{1\times F}$, initialized with small random values ($\sigma = 0.02$). For each batch, features from present views were projected normally, while absent views were filled with their corresponding learnable missing embeddings. These embeddings functioned as trainable indicators of missingness and did not attempt to simulate, reconstruct, or substitute for the unavailable view. This design prevented the model from associating zero vectors with pathological patterns and improved training stability.

When PLAX-zoomed views were present, valve features were refined using a learnable gating mechanism:

$$g = \sigma\big(\mathrm{MLP}([v, z, m_z])\big) \in \mathbb{R}^{B\times 1}$$

where $\mathrm{m}_z \in \{0,1\}^B$ indicates PLAX-zoomed view availability and $\sigma(\cdot)$ denotes the sigmoid function. The gating MLP consisted of two layers with a hidden dimension of 128 and dropout ($p = 0.1$).

The refined valve feature was computed as:

$$\tilde{v} = g \cdot z + (1 - g) \cdot v$$

When PLAX-zoomed views were absent ($\mathrm{m}_z = 0$), the gate was fixed to zero, ensuring $\tilde{\mathrm{v}} = \mathrm{v}$. This mechanism was applied identically during training and inference.

To encourage semantic alignment between PLAX-derived valve features and zoomed morphological representations, a cosine similarity–based auxiliary loss was applied for samples with available PLAX-zoomed views:

$$L_{sim} = (1 - \cos(z, v)) + \max(0, \cos(z, c) - m)$$

where $\cos(\cdot,\cdot)$ denotes cosine similarity and $m$ is a margin parameter. This loss promoted alignment between valve and zoomed features while maintaining separation from cavity representations.

**Supplementary Methods 5. Feature Fusion, Classification, and Training Objective**

Following view-specific projection and zoom-aware refinement, features from all available views were integrated through feature concatenation:

$$u = [\tilde{v}; c; a; cp; ca] \in \mathbb{R}^{B\times 5F}$$

where $[;]$ denotes concatenation, $\tilde{v}$ is the refined valve feature (obtained through zoom-aware gating when PLAX-zoomed views are available, otherwise equal to the original valve feature $v$), $c$ is the cavity feature from the PLAX view, $a$ is the chamber feature from the A4C view, and $cp$ and $ca$ denote features from PLAX and A4C color Doppler views, respectively. For views that were not available, the corresponding feature positions were filled using learnable missing embeddings, as described in **Supplementary Methods 4.**

The resulting fused representation ($5F = 1280$ dimensions, with $F = 256$) was passed to a linear classification head:

$$y_{\text{main}} = h_{\text{main}}(u) \in \mathbb{R}^{B\times K}$$

where $K = 5$ corresponds to the mitral valve etiology classes (normal, prolapse, rheumatic, functional, and degenerative). The overall training objective combined three loss components:

$$\mathcal{L} = \mathcal{L}_{\text{CE}} + \lambda_{\text{sim}} \cdot \mathcal{L}_{\text{sim}} + \lambda_{\text{gate}} \cdot \mathcal{L}_{\text{gate}}$$

where $\mathcal{L}_{\text{CE}}$ denotes the cross-entropy loss, $\mathcal{L}_{\text{sim}}$ is the cosine similarity loss applied to samples with available PLAX-zoomed views, and $\mathcal{L}_{\text{gate}}$ is an entropy-based regularization term encouraging decisive gating behavior:

$$\mathcal{L}_{\text{gate}} = -\frac{1}{B}\sum_{i=1}^{B}[g_i \log(g_i) + (1 - g_i)\log(1 - g_i)].$$

The weighting parameters were empirically set to $\lambda_{\text{sim}} = 1.0$ and $\lambda_{\text{gate}} = 3 \times 10^{-5}$ to balance classification performance and regularization.

**Supplementary Methods 6. Training Strategy**

The proposed deep learning model was optimized using the AdamW[8] optimizer with momentum parameters set to $\beta_1 = 0.9$ and $\beta_2 = 0.999$. Model training was conducted for a total of 300 epochs with a batch size of 6. To promote stable convergence and mitigate overfitting, both the learning rate and weight decay were controlled using cosine annealing schedules.[9] The learning rate schedule consisted of an initial linear warm-up phase lasting five epochs, during which the learning rate was increased linearly from zero to an initial value of $5 \times 10^{-4}$. Following the warm-up period, the learning rate was gradually decayed according to a cosine schedule to a minimum value of $1 \times 10^{-5}$ by the end of training. Weight decay was similarly governed by a cosine annealing strategy, with an initial value of 0.05 maintained throughout training. This regularization scheme was chosen to balance model generalization and learning capacity when training the multi-view architecture. Model checkpoints were evaluated at each epoch using the validation dataset, and the checkpoint achieving the best validation performance was selected for final evaluation on the internal and external test sets.

**Supplementary Results 1. Mitral Stenosis Severity Distribution in the Developmental and External Datasets**

Distribution of mitral stenosis (MS) severity (mild, moderate, and severe) across major etiologic categories in the developmental and external datasets. In contrast to mitral regurgitation (MR), the overall distribution of MS severity was largely comparable between datasets, with no consistent shift toward milder or more severe disease in the external cohort.

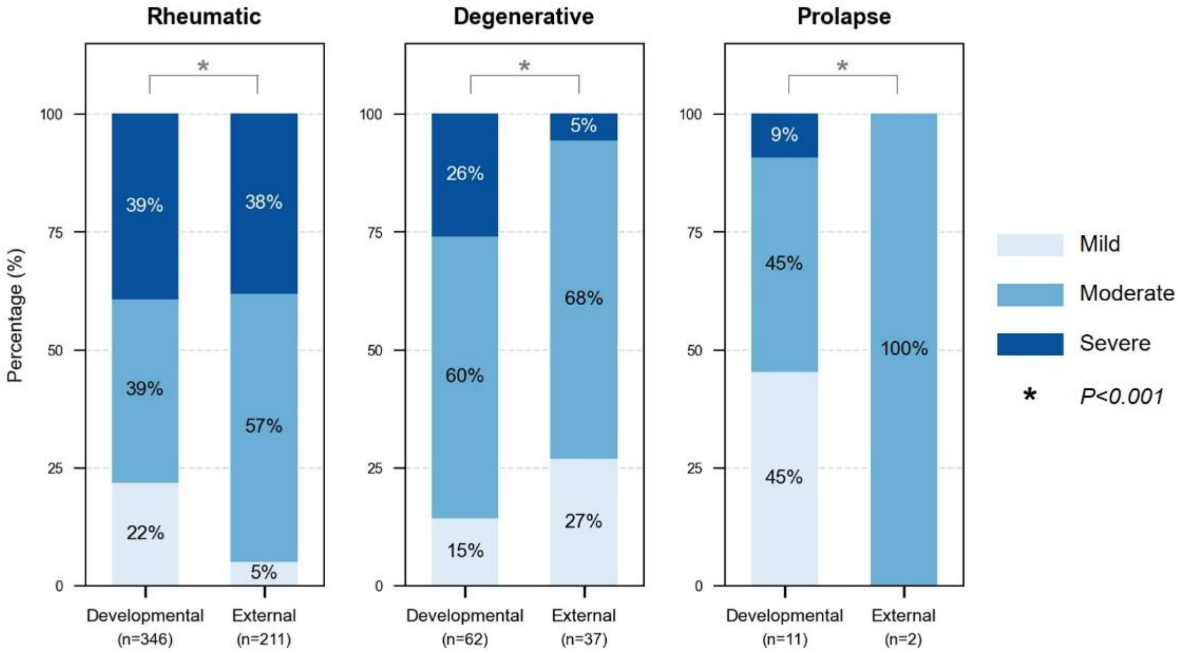

**Supplementary Results 2. Row-Normalized Confusion Matrices for MV Etiology Classification in the Internal and External Test Datasets.**

Each row was normalized to sum to 1.0, such that the displayed values represent proportion of cases within each ground-truth etiology assigned to each predicted class.

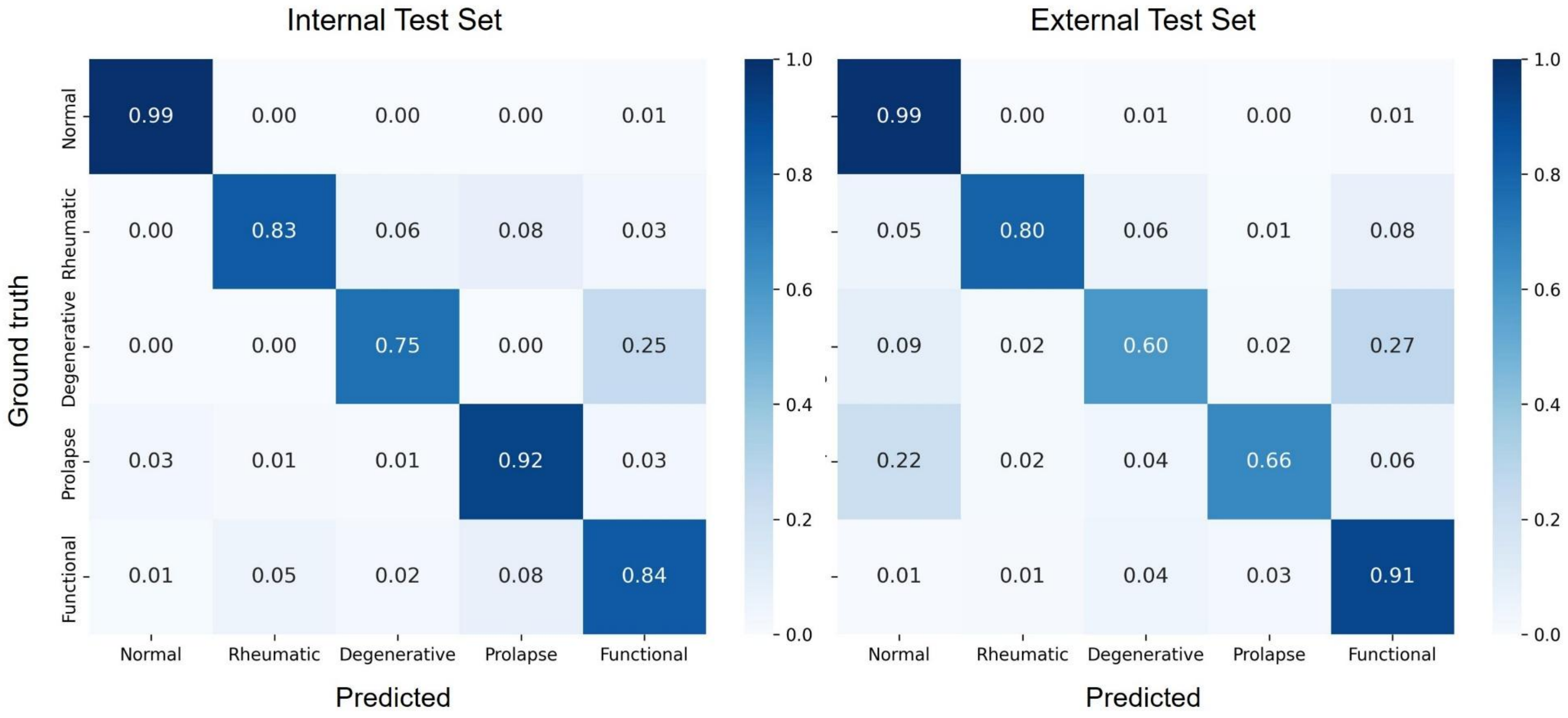

### Supplementary Results 3. UMAP-Based Characterization of Learned Feature Representations in the External Test Dataset

To further characterize the learned feature representations, additional exploratory visualization was performed using uniform manifold approximation and projection (UMAP). This analysis was conducted in the external test dataset, which provided broader representation across mitral valve (MV) etiologic categories than the internal test set. In particular, degenerative MV disease was represented by only 12 cases in the internal test set, limiting meaningful interpretation of cluster-level structure for this category.

To provide insight into how task-relevant representations emerged across the model pipeline, UMAP visualizations were generated at two stages. First, embeddings extracted from the shared MViT encoder using the parasternal long-axis (PLAX) view, which was consistently available across examinations and served as the principal morphologic input, showed substantial overlap among etiologic classes and did not demonstrate clear etiology-specific separation. Second, the final multi-view fused features, obtained after integration of refined valve, cavity, apical four-chamber, and color Doppler features, showed a more organized structure, with clearer clustering of normal and rheumatic etiologies. Residual overlap persisted among certain non-rheumatic MR etiologies, particularly functional and degenerative disease, consistent with the misclassification patterns observed in the confusion matrix and with the known morphologic and hemodynamic overlap between these categories. Partial overlap between normal and prolapse cases was also observed and may reflect milder prolapse phenotypes in the external cohort, in which morphologic abnormalities were less pronounced. Overall, these exploratory visualizations suggest that the model progressively learned more discriminative, etiology-relevant representations through multi-view fusion, while residual overlap remained in clinically challenging categories.

**UMAP-Based Visualization of Shared Encoder and Final Fused Feature Representations in the External Test Dataset.**

(A) UMAP of PLAX-based shared MViT encoder embeddings. (B) UMAP of final multi-view fused feature embeddings. Each point represents one examination and is colored by ground-truth mitral valve etiology.

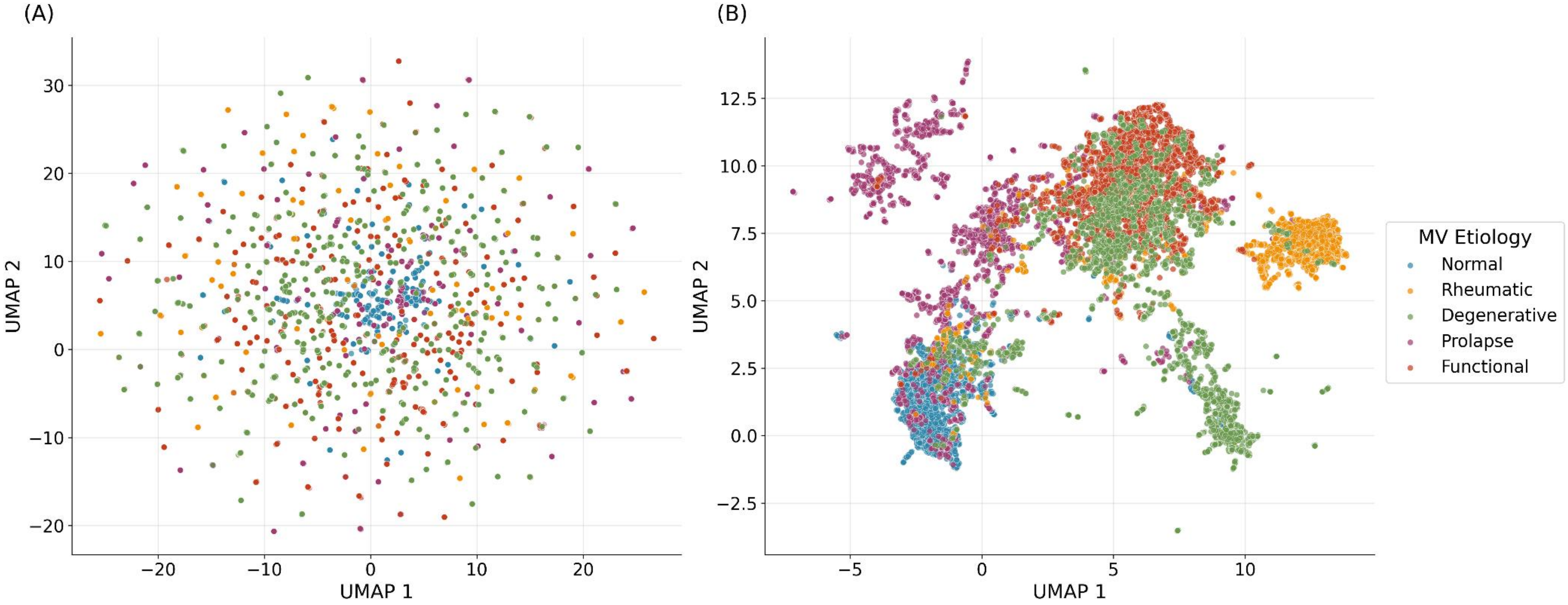

**Supplementary Results 4. MR Severity-Stratified Diagnostic Performance of the DL Model in the Internal and External Test Dataset**

To further investigate the potential impact of MR severity on etiologic classification performance, additional analyses were performed in both internal and external test datasets after restricting the cohort to cases with at least mild MR. Cases without clinically significant MR were excluded. Because these subgroup analyses excluded normal or trivial MR cases, they represent a more challenging classification setting than the full cohort and are enriched for etiologies with greater morphological overlap.

In the internal test dataset, the remaining cohort comprised 25 cases with mild MR and 247 cases with MR of moderate or greater severity. Overall accuracy was similar between the two severity strata (0.880 vs. 0.879). However, etiology-specific performance showed heterogeneous patterns across severity groups. In cases with mild MR, diagnostic performance remained high for rheumatic, degenerative, prolapse, and functional MV disease, with sensitivity of 0.833, 0.857, 1.000, and 0.909, respectively. In cases with MR of moderate or greater severity, performance also remained high for rheumatic, prolapse, and functional MV diseases, with sensitivities of 0.875, 0.921, and 0.827, respectively. In contrast, degenerative MV disease showed unstable subgroup performance, with sensitivity decreasing from 0.857 in mild MR to 0.000 in MR of moderate or greater severity. This finding should be interpreted cautiously given the very small number of degenerative cases in the latter subgroup (n=2).

In the external test dataset, the remaining cohort comprised 977 cases with mild MR and 644 cases with moderate or greater MR. Overall accuracy was higher in cases with MR of moderate or greater severity than in those with mild MR (0.856 vs. 0.676). Etiology-specific analyses again showed heterogeneous effects of MR severity. Sensitivity for prolapse increased markedly from 0.330 in mild MR to 0.837 in moderate or greater MR. In contrast, degenerative

MV disease did not show a comparable improvement, with sensitivity decreasing from 0.601 to 0.355. For rheumatic and functional MV disease, sensitivity remained relatively preserved across severity strata, changing from 0.732 to 0.797 and from 0.896 to 0.922, respectively. Confusion matrix analyses further demonstrated that misclassification in mild MR predominantly occurred among degenerative, prolapse, and functional etiologies, whereas class separation improved in cases with MR of moderate or greater severity, particularly for prolapse.

Overall, these findings suggest that the relationship between MR severity and etiologic classification performance is not uniform across datasets or etiologies. Rather, severity-stratified performance appears to reflect the combined effects of lesion severity, phenotypic overlap, subgroup size, and dataset-specific case mix.

#### *4.1. MR Severity-Stratified Diagnostic Performance of the DL Model in the Internal Test Dataset*

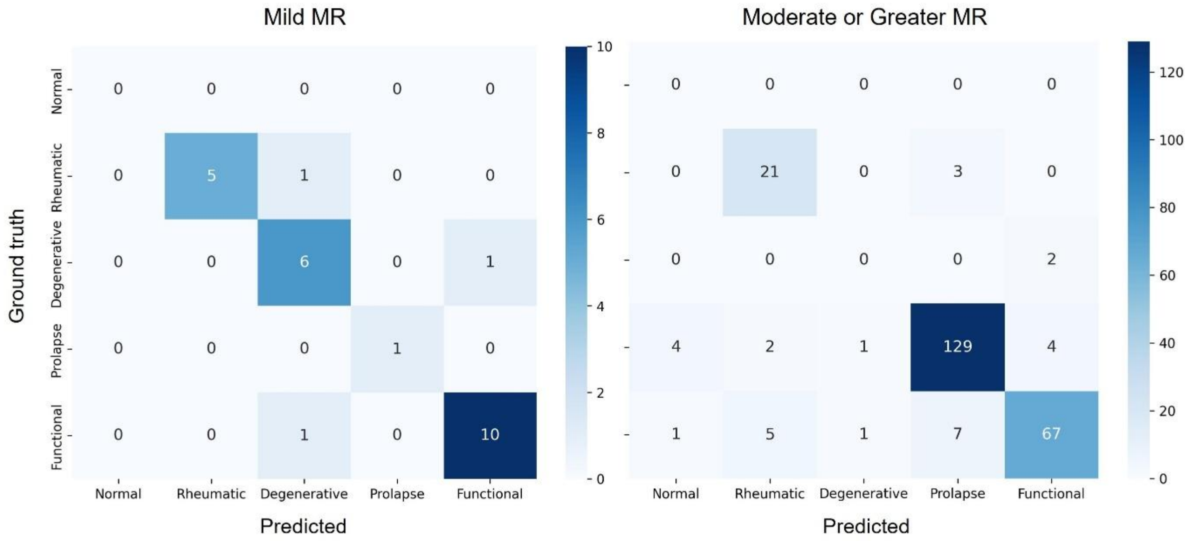


| | Mild MR | | | | | | Moderate or Greater MR | | | | | |
|---|---|---|---|---|---|---|---|---|---|---|---|---|
| | Accuracy | Precision | Sensitivity | Specificity | F1-score | AUROC | Accuracy | Precision | Sensitivity | Specificity | F1-score | AUROC |
| Normal | 1.000 | 0.000 | 0.000 | 1.000 | 0.000 | - | 0.980 | 0.000 | 0.000 | 0.980 | 0.000 | - |
| Rheumatic | 0.960 | 1.000 | 0.833 | 1.000 | 0.909 | 0.991 | 0.960 | 0.750 | 0.875 | 0.969 | 0.808 | 0.950 |
| Degenerative | 0.880 | 0.750 | 0.857 | 0.889 | 0.800 | 0.937 | 0.984 | 0.000 | 0.000 | 0.992 | 0.000 | 0.898 |
| Prolapse | 1.000 | 1.000 | 1.000 | 1.000 | 1.000 | 1.000 | 0.915 | 0.928 | 0.921 | 0.907 | 0.925 | 0.967 |
| Functional | 0.920 | 0.909 | 0.909 | 0.929 | 0.909 | 0.987 | 0.919 | 0.918 | 0.827 | 0.964 | 0.870 | 0.954 |

#### *4.2. MR Severity-Stratified Diagnostic Performance of the DL Model in the External Dataset*

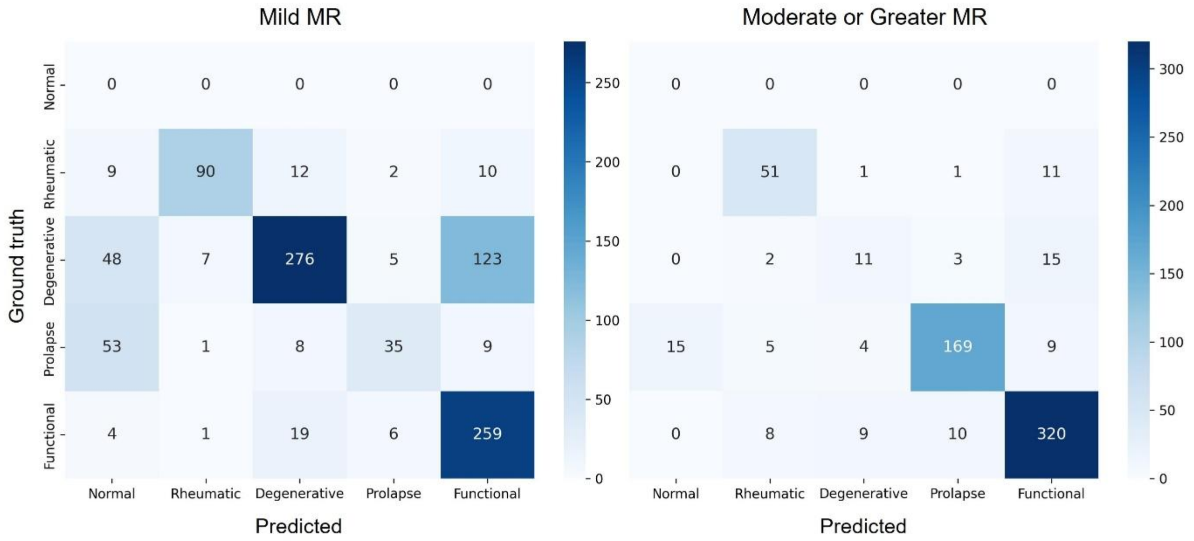


| | Mild MR | | | | | | Moderate or Greater MR | | | | | |
|---|---|---|---|---|---|---|---|---|---|---|---|---|
| | Accuracy | Precision | Sensitivity | Specificity | F1-score | AUROC | Accuracy | Precision | Sensitivity | Specificity | F1-score | AUROC |
| Normal | 0.883 | 0.000 | 0.000 | 0.883 | 0.000 | - | 0.977 | 0.000 | 0.000 | 0.977 | 0.000 | - |
| Rheumatic | 0.957 | 0.909 | 0.732 | 0.989 | 0.811 | 0.937 | 0.957 | 0.773 | 0.797 | 0.974 | 0.785 | 0.959 |
| Degenerative | 0.773 | 0.876 | 0.601 | 0.925 | 0.713 | 0.876 | 0.947 | 0.440 | 0.355 | 0.977 | 0.393 | 0.915 |
| Prolapse | 0.914 | 0.729 | 0.330 | 0.985 | 0.455 | 0.858 | 0.927 | 0.923 | 0.837 | 0.968 | 0.878 | 0.965 |
| Functional | 0.824 | 0.646 | 0.896 | 0.794 | 0.751 | 0.913 | 0.904 | 0.901 | 0.922 | 0.882 | 0.912 | 0.956 |

**Supplementary Results 5. MS Severity-Stratified Diagnostic Performance of the DL Model in the Internal and External Test Datasets**

To further explore the potential impact of mitral stenosis (MS) severity on etiologic classification performance, additional analyses were performed in the internal and external test datasets after stratifying cases into mild MS and moderate or greater MS subgroups. These analyses should be interpreted in the clinical context of MS, for which the major etiologies are predominantly rheumatic and non-rheumatic degenerative (mitral annular calcification-related) disease.

In the internal test dataset, mild MS included 9 cases and moderate or greater MS included 26 cases. In the external dataset, mild MS included 21 cases and moderate or greater MS included 229 cases. Across both datasets, the MS severity-stratified results were driven primarily by rheumatic and degenerative MV disease. Within this clinically relevant framework, diagnostic performance for rheumatic and degenerative etiologies remained generally preserved across MS severity strata.

Overall, these findings suggest that, in the setting of MS, the proposed model retained its ability to distinguish the two principal clinical etiologies across different severity levels. Because the remaining categories were uncommon in MS-stratified subgroups, these analyses should be regarded as exploratory rather than as a comprehensive multi-class severity comparison analogous to the MR analysis.

#### *5.1. MS Severity-Stratified Diagnostic Performance of the DL Model in the Internal Test Dataset*

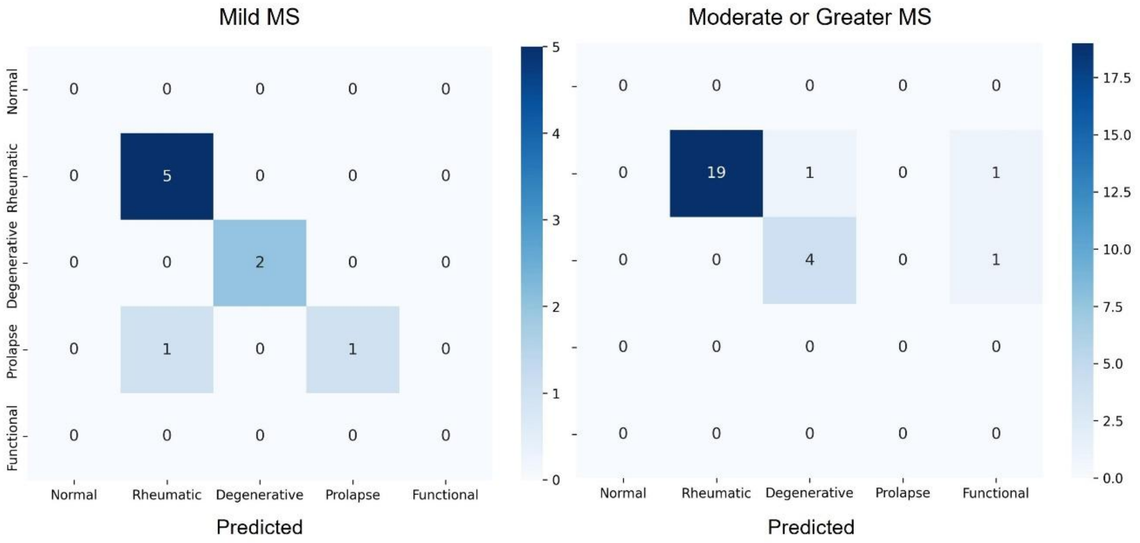


| | Mild MS | | | | | | Moderate or Greater MS | | | | | |
|---|---|---|---|---|---|---|---|---|---|---|---|---|
| | Accuracy | Precision | Sensitivity | Specificity | F1-score | AUROC | Accuracy | Precision | Sensitivity | Specificity | F1-score | AUROC |
| Normal | 1.000 | 0.000 | 0.000 | 1.000 | 0.000 | - | 1.000 | 0.000 | 0.000 | 1.000 | 0.000 | - |
| Rheumatic | 0.889 | 0.833 | 1.000 | 0.750 | 0.909 | 0.950 | 0.923 | 1.000 | 0.905 | 1.000 | 0.950 | 0.971 |
| Degenerative | 1.000 | 1.000 | 1.000 | 1.000 | 1.000 | 1.000 | 0.923 | 0.800 | 0.800 | 0.952 | 0.800 | 0.962 |
| Prolapse | 0.889 | 1.000 | 0.500 | 1.000 | 0.667 | 1.000 | 1.000 | 0.000 | 0.000 | 1.000 | 0.000 | - |
| Functional | 1.000 | 0.000 | 0.000 | 1.000 | 0.000 | - | 0.923 | 0.000 | 0.000 | 0.923 | 0.000 | - |

#### *5.2. MS Severity-Stratified Diagnostic Performance of the DL Model in the External Dataset*

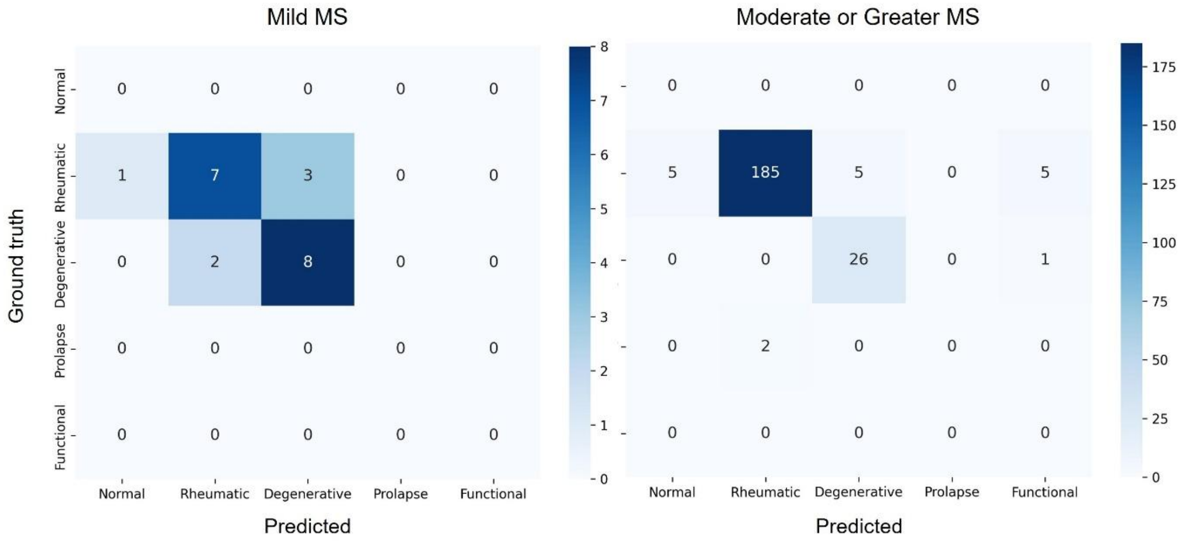


| | Mild MS | | | | | | Moderate or Greater MS | | | | | |
|---|---|---|---|---|---|---|---|---|---|---|---|---|
| | Accuracy | Precision | Sensitivity | Specificity | F1-score | AUROC | Accuracy | Precision | Sensitivity | Specificity | F1-score | AUROC |
| Normal | 0.952 | 0.000 | 0.000 | 0.952 | 0.000 | - | 0.978 | 0.000 | 0.000 | 0.978 | 0.000 | - |
| Rheumatic | 0.714 | 0.778 | 0.636 | 0.800 | 0.700 | 0.855 | 0.926 | 0.989 | 0.925 | 0.931 | 0.956 | 0.956 |
| Degenerative | 0.762 | 0.727 | 0.800 | 0.727 | 0.762 | 0.909 | 0.974 | 0.839 | 0.963 | 0.975 | 0.897 | 0.994 |
| Prolapse | 1.000 | 0.000 | 0.000 | 1.000 | 0.000 | - | 0.991 | 0.000 | 0.000 | 1.000 | 0.000 | 0.822 |
| Functional | 1.000 | 0.000 | 0.000 | 1.000 | 0.000 | - | 0.974 | 0.000 | 0.000 | 0.974 | 0.000 | - |

**Supplementary Results 6. Diagnostic Performance of the B-Mode–Only Model in the Internal and External Test Datasets**

To further evaluate the contribution of color Doppler inputs, we trained and evaluated an additional model using only B-mode echocardiographic views. Diagnostic performance of the B-mode–only model is summarized below. In the internal test dataset, the B-mode–only model showed broadly preserved diagnostic performance across etiologic categories. In the external test dataset, etiologic classification remained feasible, although performance was modestly attenuated for selected non-rheumatic MR etiologies compared with the full B-mode plus color Doppler model. In particular, degenerative MV disease showed lower sensitivity, whereas prolapse showed lower precision despite preserved sensitivity, suggesting increased false-positive prolapse classification when color Doppler information was unavailable. These findings support that B-mode views provide meaningful morphologic information for MV etiology classification, while color Doppler views contribute complementary information relevant to integrated routine echocardiographic interpretation.

**Diagnostic Performance of the B-Mode–Only Model in Internal and External Test Datasets**

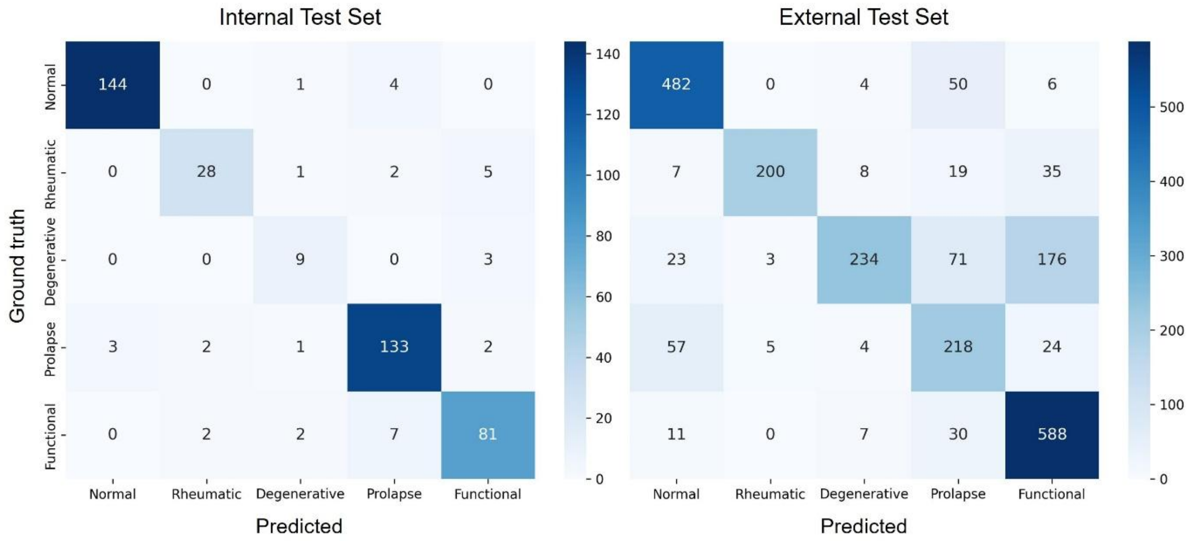


| | Internal Test Dataset | | | | | | External Test Dataset | | | | | |
|---|---|---|---|---|---|---|---|---|---|---|---|---|
| | Accuracy | Precision | Sensitivity | Specificity | F1-score | AUROC | Accuracy | Precision | Sensitivity | Specificity | F1-score | AUROC |
| Normal | 0.981 | 0.980 | 0.966 | 0.989 | 0.973 | 0.997 | 0.930 | 0.831 | 0.889 | 0.943 | 0.859 | 0.976 |
| Rheumatic | 0.972 | 0.875 | 0.778 | 0.990 | 0.824 | 0.982 | 0.966 | 0.962 | 0.743 | 0.996 | 0.839 | 0.965 |
| Degenerative | 0.981 | 0.643 | 0.750 | 0.988 | 0.692 | 0.983 | 0.869 | 0.911 | 0.462 | 0.987 | 0.613 | 0.900 |
| Prolapse | 0.951 | 0.911 | 0.943 | 0.955 | 0.927 | 0.988 | 0.885 | 0.562 | 0.708 | 0.913 | 0.626 | 0.910 |
| Functional | 0.951 | 0.890 | 0.880 | 0.970 | 0.885 | 0.977 | 0.872 | 0.709 | 0.925 | 0.852 | 0.803 | 0.946 |

### Supplementary Results 7. Distribution of Missing Input Views and Performance According to View Completeness

To further assess the robustness of the proposed framework to incomplete input availability, we analyzed the distribution of missing input views in the internal and external test datasets and compared diagnostic performance between examinations with complete views and those with one or more missing views. Detailed distributions of missing-view counts and specific missing-view combinations are summarized in the tables below.

#### 7.1. Distribution of Missing Input Views in the Internal and External Test Datasets.

| Number of missing input views | Internal test dataset, n (%) | External test dataset, n (%) |
|---|---|---|
| 0 (all required views present) | 266 (61.9%) | 2,065 (91.3%) |
| 1 | 112 (26.0%) | 170 (7.5%) |
| 2 | 32 (7.4%) | 25 (1.1%) |
| 3 | 20 (4.7%) | 2 (0.1%) |
| Total | 430 | 2,262 |

#### 7.2. Missing Input View Combinations in the Internal and External Test Datasets

| Number of Missing Views | Missing View Combination | Internal Test Dataset, n | External test Dataset, n |
|---|---|---|---|
| 1 | PLAX zoomed MV | 35 | 49 |
| 1 | A4C | 3 | 11 |
| 1 | PLAX Color Doppler | 58 | 65 |
| 1 | A4C Color Doppler | 16 | 45 |
| 2 | PLAX zoomed MV, PLAX Color Doppler | 24 | 8 |
| 2 | PLAX zoomed MV, A4C Color Doppler | 1 | 1 |
| 2 | A4C, PLAX Color Doppler | 1 | 1 |
| 2 | A4C, A4C Color Doppler | 1 | 13 |
| 2 | PLAX Color Doppler, A4C Color Doppler | 5 | 2 |
| 3 | PLAX zoomed MV, PLAX Color Doppler, A4C Color Doppler | 20 | 2 |
| Total | | 164 | 197 |

Because examinations with 2 or 3 missing views were relatively uncommon, especially in the external test dataset, performance analyses according to each exact level of missingness were considered potentially unstable. Therefore, examinations were grouped as complete-view versus incomplete-view studies. In the internal test dataset, overall accuracy, balanced accuracy, and macro F1 score were 0.895, 0.858, and 0.836 in complete-view studies, and 0.951, 0.903, and 0.912 in incomplete-view studies, respectively. In the external dataset, the corresponding values were 0.812, 0.791, and 0.802 in complete-view studies, and 0.817, 0.773, and 0.780 in incomplete-view studies, respectively.

Overall, no clinically meaningful degradation in diagnostic performance was observed in examinations with incomplete views. The numerically higher performance observed in the incomplete-view subgroup of the internal test dataset likely reflects subgroup-specific case mix rather than an advantage of missing views.

#### *7.3. Diagnostic Performance According to View Completeness in the Internal Test Dataset*

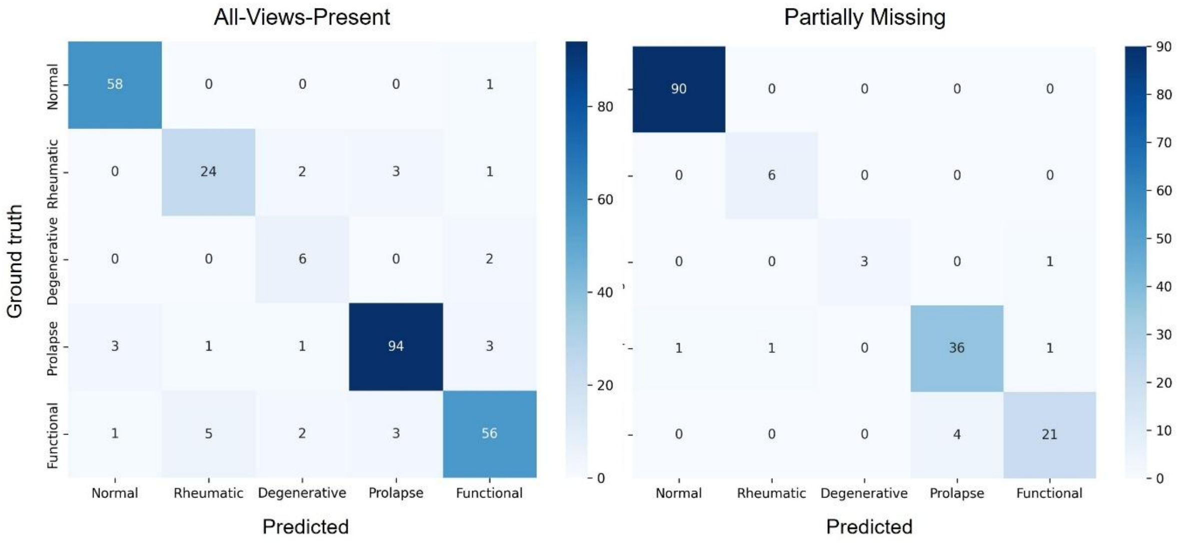


| | All-Views-Present | | | | | | Partially Missing | | | | | |
|---|---|---|---|---|---|---|---|---|---|---|---|---|
| | Accuracy | Precision | Sensitivity | Specificity | F1-score | AUROC | Accuracy | Precision | Sensitivity | Specificity | F1-score | AUROC |
| Normal | 0.981 | 0.935 | 0.983 | 0.981 | 0.959 | 0.997 | 0.994 | 0.989 | 1.000 | 0.986 | 0.994 | 1.000 |
| Rheumatic | 0.955 | 0.800 | 0.800 | 0.975 | 0.800 | 0.957 | 0.994 | 0.857 | 1.000 | 0.994 | 0.923 | 1.000 |
| Degenerative | 0.974 | 0.545 | 0.750 | 0.981 | 0.632 | 0.982 | 0.994 | 1.000 | 0.750 | 1.000 | 0.857 | 0.972 |
| Prolapse | 0.947 | 0.940 | 0.922 | 0.963 | 0.931 | 0.981 | 0.957 | 0.900 | 0.923 | 0.968 | 0.911 | 0.995 |
| Functional | 0.932 | 0.889 | 0.836 | 0.965 | 0.862 | 0.961 | 0.963 | 0.913 | 0.840 | 0.986 | 0.875 | 0.991 |

#### *7.4. Diagnostic Performance According to View Completeness in the External Dataset*

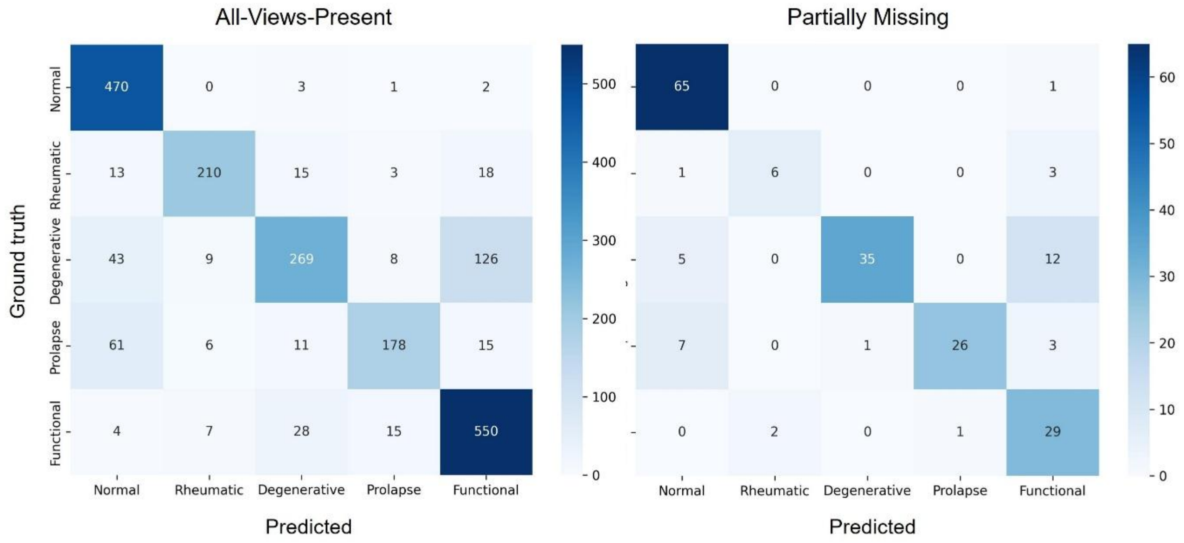


| | All-Views-Present | | | | | | Partially Missing | | | | | |
|---|---|---|---|---|---|---|---|---|---|---|---|---|
| | Accuracy | Precision | Sensitivity | Specificity | F1-score | AUROC | Accuracy | Precision | Sensitivity | Specificity | F1-score | AUROC |
| Normal | 0.938 | 0.795 | 0.987 | 0.924 | 0.881 | 0.993 | 0.929 | 0.833 | 0.985 | 0.901 | 0.903 | 0.988 |
| Rheumatic | 0.966 | 0.905 | 0.811 | 0.988 | 0.855 | 0.971 | 0.970 | 0.750 | 0.600 | 0.989 | 0.667 | 0.841 |
| Degenerative | 0.882 | 0.825 | 0.591 | 0.965 | 0.689 | 0.928 | 0.909 | 0.972 | 0.673 | 0.993 | 0.795 | 0.964 |
| Prolapse | 0.942 | 0.868 | 0.657 | 0.985 | 0.748 | 0.942 | 0.939 | 0.963 | 0.703 | 0.994 | 0.812 | 0.944 |
| Functional | 0.896 | 0.774 | 0.911 | 0.890 | 0.837 | 0.957 | 0.888 | 0.604 | 0.906 | 0.885 | 0.725 | 0.949 |

**Supplementary Results 8. Distribution and Model Performance in Cases with Multiple Mitral Valve Etiologies**

A total of 63 cases were identified as having multiple coexisting mitral valve (MV) etiologies based on expert review. The distribution of coexisting etiologic combinations is illustrated in a pie chart, demonstrating the relative frequency of each combination.

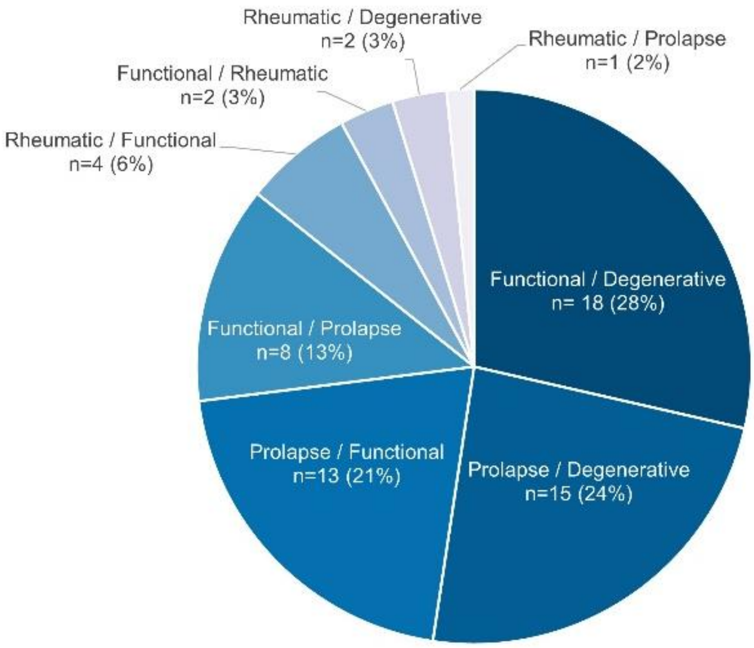


Although these cases were excluded from model training, post-hoc inference was performed to evaluate model behavior in this clinically relevant subgroup. For each case, model predictions were categorized as correctly identifying the primary etiology, correctly identifying the secondary etiology, or being misclassified into an etiologic category not assigned by expert review.

| Primary Etiology | Secondary Etiology | N | Primary Correct | Secondary Correct | Misclassified |
|---|---|---|---|---|---|
| Functional | Degenerative | 18 | 12 (66.7%) | 0 (0.0%) | 6 |
| Prolapse | Degenerative | 15 | 13 (86.7%) | 0 (0.0%) | 2 |
| Prolapse | Functional | 13 | 10 (76.9%) | 2 (15.4%) | 1 |
| Functional | Prolapse | 8 | 3 (37.5%) | 5 (62.5%) | 0 |
| Rheumatic | Functional | 4 | 1 (25.0%) | 3 (75.0%) | 0 |
| Functional | Rheumatic | 2 | 0 (0.0%) | 2 (100%) | 0 |
| Rheumatic | Degenerative | 2 | 2 (100%) | 0 (0.0%) | 0 |
| Rheumatic | Prolapse | 1 | 1 (100%) | 0 (0.0%) | 0 |
| Total | | 63 | 42 (66.7%) | 12 (19.0%) | 9 |

**Supplementary Results 9. Image Quality-Stratified Performance Analysis.**

Image quality (IQ)–stratified analyses were performed to evaluate the robustness of the proposed DL model under varying imaging conditions. Examinations were initially categorized into three predefined IQ strata based on automated assessment of B-mode views: High, in which all input views were of at least fair quality; Mixed, in which fair or higher–quality views coexisted with poor or non-diagnostic views; and Low, in which all available views were classified as poor or non-diagnostic. However, across both test datasets, the number of IQ Low examinations was extremely limited (internal: n = 1; external: n = 6). Given this marked imbalance, the Mixed and IQ Low categories were combined into a single group, referred to as Partially Suboptimal, to ensure stable and interpretable subgroup comparisons. Accordingly, subsequent analyses compared All-Adequate (IQ High) versus Partially Suboptimal (Mixed + IQ Low) examinations.

**Distribution of IQ Categories**

| | IQ | Internal | External |
|---|---|---|---|
| All-Adequate | High | 293 | 1691 |
| Partially Suboptimal | Mixed | 136 | 565 |
| | Low | 1 | 6 |

Detailed analyses stratified by IQ in the internal and external test datasets are as follows.

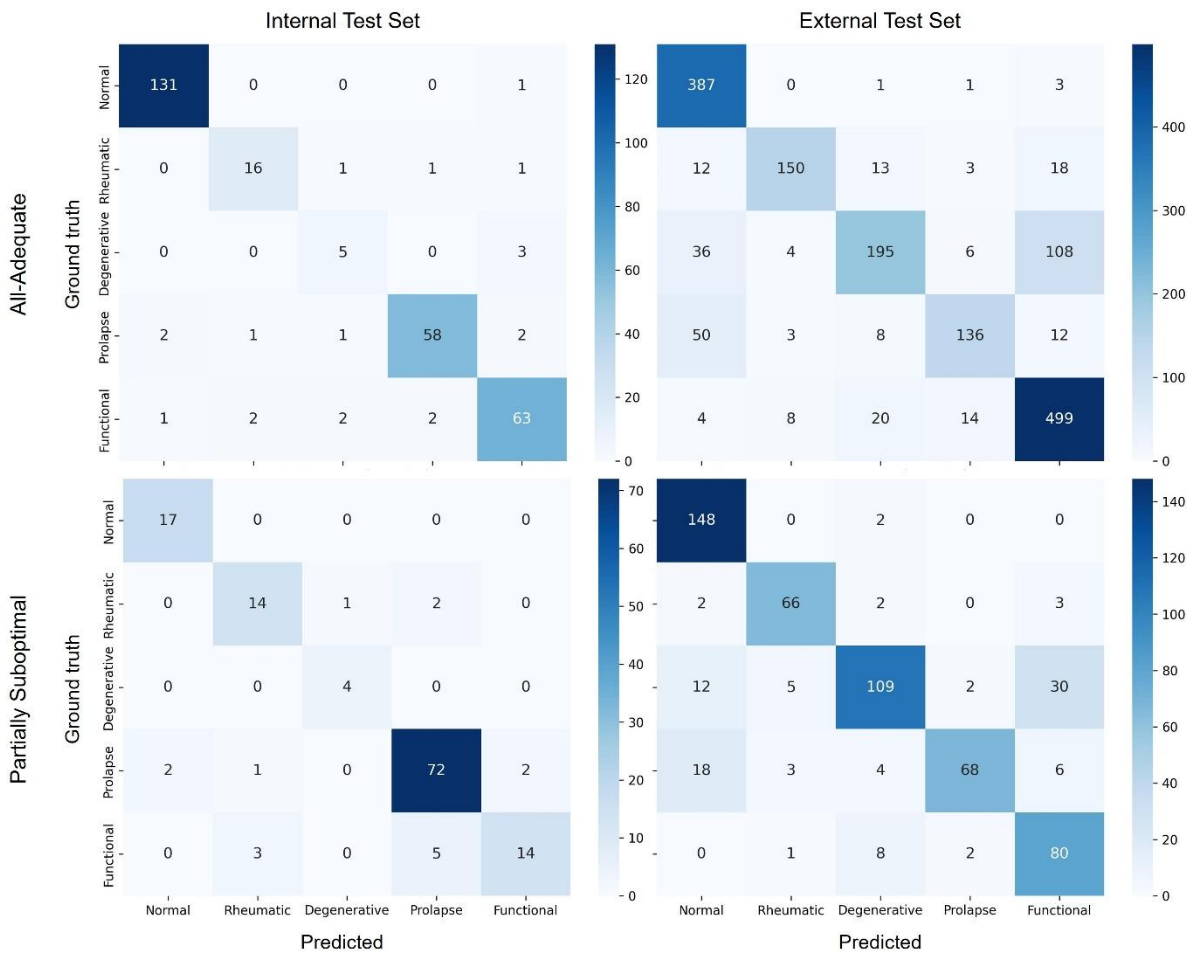

**Internal Test Dataset**

| | All-Adequate | | | | | | Partially Suboptimal | | | | | |
|---|---|---|---|---|---|---|---|---|---|---|---|---|
| | Accuracy | Precision | Sensitivity | Specificity | F1-score | AUROC | Accuracy | Precision | Sensitivity | Specificity | F1-score | AUROC |
| Normal | 0.986 | 0.978 | 0.992 | 0.981 | 0.985 | 0.996 | 0.985 | 0.895 | 1.000 | 0.983 | 0.944 | 0.996 |
| Rheumatic | 0.980 | 0.842 | 0.842 | 0.989 | 0.842 | 0.983 | 0.949 | 0.778 | 0.824 | 0.967 | 0.800 | 0.940 |
| Degenerative | 0.976 | 0.556 | 0.625 | 0.986 | 0.588 | 0.970 | 0.993 | 0.800 | 1.000 | 0.992 | 0.889 | 1.000 |
| Prolapse | 0.969 | 0.951 | 0.906 | 0.987 | 0.928 | 0.988 | 0.912 | 0.911 | 0.935 | 0.883 | 0.923 | 0.966 |
| Functional | 0.952 | 0.900 | 0.900 | 0.969 | 0.900 | 0.980 | 0.927 | 0.875 | 0.636 | 0.983 | 0.737 | 0.925 |

**External Test Dataset**

| | All-Adequate | | | | | | Partially Suboptimal | | | | | |
|---|---|---|---|---|---|---|---|---|---|---|---|---|
| | Accuracy | Precision | Sensitivity | Specificity | F1-score | AUROC | Accuracy | Precision | Sensitivity | Specificity | F1-score | AUROC |
| Normal | 0.937 | 0.791 | 0.987 | 0.921 | 0.879 | 0.994 | 0.940 | 0.822 | 0.987 | 0.924 | 0.897 | 0.986 |
| Rheumatic | 0.964 | 0.909 | 0.765 | 0.990 | 0.831 | 0.962 | 0.972 | 0.880 | 0.904 | 0.982 | 0.892 | 0.979 |
| Degenerative | 0.884 | 0.823 | 0.559 | 0.969 | 0.666 | 0.927 | 0.886 | 0.872 | 0.690 | 0.961 | 0.770 | 0.941 |
| Prolapse | 0.943 | 0.850 | 0.651 | 0.984 | 0.737 | 0.936 | 0.939 | 0.944 | 0.687 | 0.992 | 0.795 | 0.955 |
| Functional | 0.889 | 0.780 | 0.916 | 0.877 | 0.842 | 0.953 | 0.912 | 0.672 | 0.879 | 0.919 | 0.762 | 0.966 |
